# Multivariate genome-wide association study of suicidal behaviors in >1.7 million individuals of diverse population descents

**DOI:** 10.64898/2025.12.15.25342298

**Authors:** Jun He, Brenda Cabrera-Mendoza, Dan Qiu, David Davtian, Zhongzheng Mao, Qianyu Chen, Eric N. Penichet, Qishu Zhang, Sefayet Karaca, Renato Polimanti

**Affiliations:** Department of Psychiatry, Yale School of Medicine, New Haven, CT, USA; Veteran Affairs Connecticut Healthcare System, West Haven, CT, USA; University of Connecticut, Storrs, CT, USA; Center for Otolaryngology and Hearing Care, Jinan Maternity and Child Care Hospital Affiliated to Shandong First Medical University, Jinan, Shandong, China; Faculty of Health Science, Aksaray University, Aksaray, Turkiye; Department of Chronic Disease Epidemiology, Yale School of Public Health, New Haven, CT, USA; Wu Tsai Institute, Yale University, New Haven, CT, USA; Department of Biomedical Informatics and Data Science, Yale School of Medicine, New Haven, CT, USA

## Abstract

**Background:** While previous genome-wide association studies (GWAS) identified multiple risk loci for suicide ideation (SI) and suicide attempt (SA), there is still a limited understanding of the genetic predisposition underlying suicidal behaviors in diverse populations. This study aimed to conduct a large-scale investigation of the suicidality spectrum (SP) to generate new insights into its biology and epidemiology.

**Methods:** Leveraging ancestrally diverse participants (SI N_case/control_=179 881/1 013 900; SA N_case/control_=66 867/1 654 798) from the UK Biobank, All of Us Research Program, Million Veteran Program, FinnGen, and Psychiatric Genomics Consortium, we performed GWAS meta-analyses for SI and SA, and a multivariate GWAS of SP. We applied multiple analytical approaches to identify genomic loci associated with suicide traits and to functionally annotate these significant signals. Phenome-wide genetic correlation and genetically informed causal analyses were further conducted to provide convergent evidence on the relationships of suicidal behaviors with health outcomes, brain imaging-derived phenotypes, and metabolomic traits.

We identified 90 independent lead single-nucleotide polymorphisms (SNPs) for suicidal behaviors, of which 49 were novel. SNP-based heritability was higher for SA (SNP-h^2^=0.115±0.005) than for SI (SNP-h^2^=0.040±0.002) and SP (SNP-h^2^=0.050±0.002), and their genetic correlations ranged from 0.639 to 0.960. Functional annotation analyses highlighted associated genes (e.g., *UGGT2*, *GMPPB*, *BRWD1*), enriched gene sets (e.g., cellular response to stress), and potential therapeutic drug candidates (e.g., cariprazine, paliperidone, droperidol), with some signals shared across suicide traits but more specific to SI or SA. Suicide behaviors were genetically associated with a broad spectrum of complex traits, primarily in the domains of mental health, physical health, behaviors, and socioeconomic factors, some of which revealed causal relationships.

**Interpretation:** This study provides convergent genetic evidence for both shared and phenotype-specific components of suicidal behaviors and delineates their associated factors spanning from proximal clinical and behavioral traits to more distal social determinants. These findings refine our understanding of the etiology of suicidal behaviors and may inform targeted strategies for suicide prevention in both clinical and public health settings.

**Research in context:** *Evidence before this study:* Suicidal behaviors remain a serious public health concern and contribute to substantial mortality globally. However, their genetic predisposition and risk profiles associated with them are not yet fully established although multiple approaches, including genome-wide association studies (GWAS), have been applied. We searched PubMed, medRxiv, and bioRxiv for publications and preprints in English from Jan 1, 2000, to Nov 1, 2025, using the search terms “suicid*” and “GWAS”. The largest previously published GWASs identified four genome-wide loci for suicide ideation and 12 for suicide attempt, and a recent preprint reported 77 loci for suicidal behaviors. These associations were mainly derived from individuals of European ancestry, with only one significant locus for suicide ideation reported in East Asian ancestry. Previous studies also attempted to prioritize putative risk genes, but most did not leverage multi-omic approaches, such as transcriptome-wide and proteome-wide association studies. Moreover, earlier work investigated the relationships between suicidal behaviors and psychiatric and behavioral traits mainly through genetic correlation analysis, without conducting phenome-wide genetically informed causal analysis.

*Added value of this study:* Our study identified 90 genome-wide significant associations for suicidality. These associations were distributed across European, African, Admixed American, and Asian ancestries for both suicide ideation and suicide attempt. SNP-based heritability for suicidal behaviors ranged from 4% to 12% and remained significant after conditioning on major psychiatric disorders. We prioritized 1052 genes associated with suicidal behaviors and identified 33 loci with convergent evidence across five or more gene-discovery analyses. Sixteen genes were shared across suicide ideation, suicide attempt, and suicidality spectrum, while most other loci appeared to be phenotype-specific. Drug-repurposing analysis suggested five potential therapeutic drug candidates: cariprazine, droperidol, molindone, paliperidone, and chlorprothixene. Using phenome-wide genetically informed analyses, we identified loneliness, medical abortion, and age at first sexual intercourse as putative causal risk factors for suicidal behaviors. Suicidal behaviors were also associated with adverse consequences, including hospital admissions and multiple mental health and physical conditions.

*Implications of all the available evidence:* The predisposition to suicidal behaviors is due to genetic mechanisms acting through molecular changes across multiple omic domains in brain and peripheral systems. Although psychiatric disorders may mediate the genetic liability of suicidal behaviors, a proportion of the risk appears to be independent of psychiatric diagnosis. Suicidal behaviors are influenced by diverse genetic and causal risk factors, and can also lead to broad adverse health consequences.

## Introduction

Suicide remains a major global public health concern. In 2023, an estimated 766 700 individuals died by suicide worldwide, corresponding to an age-standardized mortality rate of 9.0 per 100 000 population.^1^ Although the global rate has been declining over decades, suicide incidence was still increasing in several regions, including Latin America, North America, and Asia–Pacific region.^1,2^ Regarding age-specific observations, suicide was the third leading cause of death among adolescents and young adults aged 10–29 years,^3^ and suicide rate was highest among adults aged 70 years and over.^4^ Suicidal behaviors, including suicide ideation (SI) and suicide attempt (SA), are well-established predictors of suicide death.^5^ These suicidal traits are related to a range of adverse outcomes, including long-term mental and physical health complications,^6^ imposing a substantial economic burden on the health system and society.^7,8^

Previously published genome-wide association studies (GWAS) have identified a limited number of significant loci associated with suicide traits, primarily due to modest sample sizes.^9–19^ More recently, a large-scale GWAS meta-analysis reported 59 previously unreported genomic loci for suicidality, offering novel biological insights.^20^ Despite the progress, most previous studies have concentrated on variant-level associations, with relatively few efforts directed toward comprehensive gene-level prioritization. In addition, most genetic findings have been derived from individuals of European (EUR) ancestry, providing limited evidence for other ancestral groups. For example, although a GWAS reported genome-wide significant loci for SA in African (AFR), Admixed American (AMR), and Asian (ASN) descents,^14^ the largest previous GWAS identified only one single-nucleotide polymorphism (SNP) for SI in East Asian (EAS) ancestry.^20^ Existing GWASs have predominantly focused on relationships between suicide traits and major psychiatric disorders, providing limited exploration across a broader range of associated factors. Multiple risk factors have been implicated in observational studies, including genetic, clinical, psychosocial, and environmental aspects.^21–23^ However, most previous studies have typically examined narrow dimensions based on ecological, case–control, or psychological-autopsy designs that primarily suggested correlation-level findings, with limited evidence from causally informative analyses to identify broader etiologically meaningful factors and consequences of suicide.^24,25^ Given the rapid expansion of genomic research and the growing availability of large-scale biobank datasets, genetically informed analyses are offering powerful “natural experiments” that can be leveraged to strengthen causal inference in observational settings, providing an opportunity to move beyond correlation and more rigorously identify modifiable risks and health consequences for suicide traits, while reducing susceptibility to confounding bias inherent in traditional observational study designs.^26^

Here, leveraging data from more than 1.7 million individuals available from the UK Biobank (UKB),^27^ All of Us Research Program (AoU),^28^ Million Veteran Program (MVP),^29^ FinnGen,^30^ and Psychiatric Genomics Consortium (PGC),^31,32^ we conducted a multi-ancestry GWAS meta-analysis to identify genomic loci associated with SI and SA, and performed a multivariate GWAS of suicidality spectrum (SP). Integrating multi-omics approaches into our gene-discovery investigation, we prioritized high-confidence risk loci and biological pathways and translated them into therapeutic candidates for suicidal behaviors. We further investigated bi-directional relationships between suicide phenotypes and multiple domains, uncovering potential causal effects convergent across several genetically informed frameworks. Overall, this study provides a comprehensive characterization of the genetic architecture of suicidality, offering insights into the mechanisms underlying the occurrence and progression of suicidal behaviors.

## Methods

### Study design

We conducted GWAS for SI and SA in diverse ancestries available in UKB^27^ and AoU.^28^ Leveraging these results together with previously published GWASs from MVP,^29^ FinnGen,^30^ and PGC,^31,32^ we used genomic structural equation modeling (gSEM)^33^ to integrate EUR GWASs for SI, SA, and SP (jointly modeling SI and SA) separately. For non-EUR descent, we performed ancestry-specific meta-analysis using METAL,^34^ followed by a cross-ancestry meta-analysis combining the EUR gSEM results with non-EUR GWASs to improve generalizability across populations. Lead SNPs were identified using Functional Mapping and Annotation (FUMA).^35^ To explore secondary independent signals, we applied a stepwise model with conditional and joint association analysis (COJO)^36^, and performed fine-mapping with SuSiEx^37^ to pinpoint putative causal variants. The second part of this study consisted of comprehensive functional annotation analyses for suicide traits, including MAGMA gene-based analysis,^38^ transcriptome-wide association studies (TWAS),^39,40^ isoform-level TWAS (isoTWAS),^41^ proteome-wide association studies (PWAS),^42^ and methylation quantitative trait loci based summary-data Mendelian randomization (mQTL-SMR).^43,44^ Genes were prioritized based on the number of analyses providing convergent evidence. We further applied MAGMA gene expression analysis^38^ to identify tissues enriched for GWAS signals, used GSA-MiXeR^45^ to detect enriched gene ontologies (GO), and conducted drug repurposing analysis using DRUGSETS^46^ to identify potential therapeutic drug candidates for suicidal behaviors. In the third part of phenome-wide analyses, we first estimated genetic correlations and the proportion of shared causal variants between suicide traits and six major psychiatric disorders using linkage disequilibrium score regression (LDSC)^47,48^ and MiXeR.^49^ We subsequently extended LDSC analysis to more than 10,000 complex traits. For those showing significant genetic correlations, we applied latent causal variable model (LCV),^50^ MRlap,^51^ and generalized summary-data Mendelian randomization (GSMR2)^52^ to evaluate potential bidirectional causal relationships with suicide traits.

### Participants and GWAS data

In total, we included 1 193 781 individuals (N_case/control_=179 881/1 013 900) from UKB,^27^ AoU,^53^ and MVP^29^ for SI, and 1 721 665 individuals (N_case/control_=66 867/1 654 798) from UKB,^27^ AoU,^53^ FinnGen,^30^ and PGC^17^ for SA (appendix pp 1 Table S1A). These samples were distributed across EUR, AFR, AMR, EAS, Central/South Asian (CSA), and Middle Eastern (MID) ancestries, after restricting analyses to population groups with more than 100 cases per cohort. In the SI GWAS meta-analysis, to maintain consistency with the MVP protocol,^16^ we combined EAS and CSA into one ASN ancestry group. All cohorts received approval from their local Institutional Review Boards (IRBs), and the present study was determined by the Yale IRB to not require review and approval (Protocol ID: 2000038313) because it does not constitute human subject research.

UKB is a large population-based cohort collecting genetic and health-related data from over 500,000 individuals aged 37 to 73 years in the United Kingdom.^27^ We used electronic health records (EHR) and self-reported information to define suicide phenotypes (appendix pp 1 Table S1B). For suicide ideation analysis, individuals with a history of SA were excluded to prevent phenotype overlapping. Participants with SA, suicide death, or self-harm accompanied by any SI were categorized as SA cases. SI cases were also excluded from SA controls to ensure case-control independence. In UKB, genome-wide genotyping was performed using the UKB Axiom Array and imputed using the Haplotype Reference Consortium and UK10K+1000 Genomes reference panels. Detailed genotyping and quality-control (QC) pipelines have been reported elsewhere.^27^ We applied additional QC with criteria of minor allele frequency (MAF)>1%, Hardy–Weinberg equilibrium *P*>1×10^-6^, imputation INFO>0.8, variant missing call rate<5%, and sample missing rate<5%. Genetically inferred ancestry groups and unrelated participants were defined from the Pan-UKB study.^54^ We performed GWAS for SI (N_case/control_=52 065/216 922) in individuals of EUR, AFR, AMR, EAS, CSA, and MID descent and SA (N_case/control_=6143/305 379) in EUR using logistic regression with Firth correction in PLINK 2,^55^ with sex, age, the top 10 within-ancestry principal components (PCs) as covariates.

AoU is a nationwide, ancestrally diverse cohort in the United States that has recruited more than 633,000 participants aged 18 years or older to date.^28^ Suicide traits were defined using EHR and self-reported information (appendix pp 1 Table S1B). Genetic information was derived from whole-genome sequencing data using the ACAF threshold callset, with QC and ancestry-inference procedures described previously.^53^ Consistent with GWAS in UKB, we applied the same process for suicide-phenotype definition and additional genotype QC. GWAS for SI (N_case/control_=28 002/284 411) in EUR, AFR, AMR, EAS, and CSA descent and SA (N_case/control_=7748/280 350) in EUR, AFR, AMR, and EAS descent was conducted using logistic regression with Firth correction in PLINK 2, adjusting for sex, age, and the top 10 within-ancestry PCs.

We also included GWAS data for suicide traits from previously published studies. MVP is an observational cohort of the United States Department of Veterans Affairs (VA) healthcare system with over one million racially/ethnically diverse participants enrolled to date.^29^ GWAS for SI (defined from EHR data; without SA history) was previously conducted in participants of EUR, AFR, AMR, and ASN ancestry (N_case/control_=99 814/512 567), adjusting for sex, age and genetic substructure.^16^ The previously reported MVP SA GWAS was included in the PGC study.^14^ FinnGen is a research initiative in Finland that utilizes imputed genotype data from Finnish biobanks and digital healthcare information from national health registers.^30^ We obtained the GWAS data for SA in EUR descent (N_case/control_=11 538/488 810) from its release 12, where SA was defined as suicide or other intentional self-harm with ICD code (ICD-10: X60–X84, ICD-9: E95, or ICD-8: E95). PGC is a global collaborative program with the goal of advancing genetic discovery of biologically, clinically, and therapeutically meaningful insights of psychiatric disorders.^31,32^ In its publicly available GWAS meta-analysis of SA, 22 cohorts with multiple case definition methods (i.e., EHR, interviews, and self-/coroner’s reports) were integrated with individuals from EUR, EAS, AFR, and AMR. For the present study, we used the ancestry-specific GWAS results (N_case/control_=41 438/580 259), excluding UKB samples.^17^

### GWAS meta-analysis

For EUR ancestry, due to the relatively low genetic correlations among the cohorts investigated (median SI r_g_=0.554, median SA r_g_=0.740, and median SI–SA r_g_=0.563; appendix pp 3 Figure S1; appendix pp 1 Table S2B), we combined GWASs for SI (UKB, AoU, and MVP) and SA (UKB, AoU, FinnGen, and PGC) using gSEM.^33^ Considering SI and SA as different stages of suicidality, we also combined the EUR SI and SA GWASs as SP. Parallel analysis with the paLDSC function was performed to determine the number of latent factors.^56^ As recommended by gSEM developers, the loading of the first GWAS was fixed to 1 for model identification. Since gSEM smooths the non-positive-definite covariance matrix and calculates Z_smooth_ for the largest difference between pre- and post-smoothed matrices, we retained only variants with Z_smooth_<0.025 and *P*_Q_ (heterogeneity test)≥0.01 in the results. For non-EUR ancestries, due to limited sample sizes and lower genetic correlations, we used METAL^34^ to perform ancestry-specific, sample-size-weighted meta-analysis for SI in AFR, AMR, and ASN, and for SA in AFR, AMR, and EAS ancestries. Finally, using the EUR results from gSEM together with non-EUR GWASs, we performed a cross-ancestry meta-analysis to combine all available ancestries to improve statistical power. Only variants with MAF>1%, a total sample size>10 000, and presence in multiple cohorts (≥2 for ancestry-specific and ≥3 for cross-ancestry analyses) or with *P*<0.05 were included in the meta-analysis.

We used linkage disequilibrium score regression (LDSC) to estimate SNP-based heritability of suicide traits (both cohort-specific and combined) and their genetic correlations.^47,48^ LD scores were calculated using HapMap 3 variants^57^ and the 1000 Genomes Project Phase 3 data^58^ in the corresponding populations. SNP-based heritability for binary suicide traits was estimated on the liability scale using population prevalences of SI (9%)^16^ and SA (2%)^17^, while heritability for suicide latent factors was estimated on the observed scale using effective sample sizes.^47^ A nominal threshold was applied to identify significant heritability, and Bonferroni correction was used to account for multiple tests in estimating genetic correlations.

### Genomic loci identification

Genomic loci and lead SNPs in gSEM/METAL-combined GWASs were identified using FUMA web-based platform,^35^ with the parameters of *P*_GWAS_<5×10^-8^, LD r^2^<0.1 (LD r^2^<0.6 for independent significant SNPs), and a maximum of 250 kb window to merge genomic loci. The LD threshold of r^2^<0.1 was used to define independent lead SNPs across ancestries and suicide traits. In addition to LD-based filtering (r^2^<0.1), we also applied a ±1Mb window to determine novel SNPs more stringently by comparing with previously reported independent SNPs associated with suicidality.^9–20^ Complex traits in the GWAS Catalog^59^ previously associated with the lead SNPs or SNPs in LD with them were reported.

GCTA-COJO joint analysis^36,60^ was performed to identify secondary association signals for gSEM derived suicide traits in EUR ancestry. A stepwise model selection procedure was applied to select significant SNPs after conditioning analysis. UKB EUR genotype data from 5000 randomly sampled individuals were used as the LD reference panel, and the default parameters of GCTA-COJO were adopted. We reported SNPs with genome-wide significance (P_J_<5×10^−8^) and independence of the lead SNPs identified by FUMA (LD r^2^<0.1).

To identify putative causal variants underlying suicide traits, we performed multiple-ancestry fine-mapping analysis using SuSiEx.^37^ Combined GWASs of ancestries showing significant associations (*P*<5×10^-8^) of SI (EUR, AFR, AMR, and ASN), SA (EUR, AFR, AMR, and EAS), and SP (EUR) were considered. The 1000 Genomes Project Phase 3 data^58^ was used as reference panels to calculate LD matrices in the corresponding populations. All parameters in SuSiEx were set to their default values. Fine-mapping was conducted on the significant genomic regions determined by FUMA and 95%-credible sets were identified by SuSiEx. Variants with a maximum posterior inclusion probability (PIP)≥0.8 within these 95%-credible sets were considered putative causal variants. Variants identified in GCTA-COJO and fine-mapping analyses were mapped to genes using the Variant Effect Predictor (VEP) tool.^61^

### Functional annotation

We performed MAGMA (v1.10) gene-based and gene expression analyses,^38^ which were integrated into the FUMA web-based platform for GWASs across all available ancestries. The gene-based analysis annotated GWAS variants to the gene level, and the gene expression analysis further evaluated gene expression enrichment in 53 tissue types from GTEx v8,^62^ using the 1000 Genomes Project Phase 3 data in each ancestry as the LD reference panel,^58^ and the maximum *P*-value cutoff for annotation as 0.05. Bonferroni correction was used to account for the number of genes and tissue types tested.

Tissue-specific and cross-tissue TWAS of gSEM derived suicide traits in EUR ancestry were conducted to identify gene associations with suicidality using S-PrediXcan^39^ and S-MultiXcan.^40^ Predictive models of gene expression and their corresponding LD covariance matrices were built based on GTEx v8 data.^39,63^ We estimated tissue-specific and cross-tissue associations across 13 brain tissues. Subsequently, we performed tissue-specific analysis across 36 (non-brain) body tissues, followed by a cross-tissue analysis integrating all 49 GTEx tissues. Bonferroni correction was applied to adjust the number of genes tested for each suicide trait.

We further utilized a complementary method, isoTWAS, to investigate the associations between gSEM derived suicide traits and predicted gene expression at isoform-level resolution.^41^ Precomputed expression weights for each isoform and matched-LD matrices were derived from the PsychENCODE adult frontal cortex dataset by the developers.^64^ First, a burden test was applied to aggregate SNP effects per isoform, followed by a permutation test to generate a null distribution of SNP-isoform effects. To account for multiple testing, isoform-level *P*-values within each gene were collated to produce a gene-level screening *P*-value, followed by a false discovery rate (FDR) correction. For genes with FDR q<0.05, a within-gene significance threshold (α_2_) was computed based on the total number of filtered genes and tested genes. Isoforms were retained if they met the following criteria: (1) gene-level q-value<0.05, (2) within-gene *P*-value<α₂, and (3) permutation *P*-value<0.05. Fine-mapping was performed for genes with two or more significant isoforms, and isoforms with PIP≥0.8 were retained in the credible sets. For genes with only one significant isoform, the isoform was assigned a PIP of 1 and included in the credible set.

PWAS was conducted using FUSION^42^ to identify genes with cis-regulated protein abundance in the dorsolateral prefrontal cortex (DLPFC) associated with gSEM derived suicide traits in EUR ancestry. Following the developers’ recommendations, we employed elastic-net models due to their superior predictive performance relative to other modeling approaches. cis-pQTL weights were trained on postmortem brain tissue from two datasets: the Religious Orders Study and Memory and Aging Project (ROSMAP; N=330)^65^ and the Banner Sun Health Research Institute (Banner; N=149).^66^ These models collectively cover 7376 brain proteins measured in individuals of European ancestry. We consequently performed a sample-size-weighted meta-analysis using METAL^34^ to pool protein effects from the two datasets. The European reference panel from the 1000 Genomes Project^58^ was used for LD estimation. To adjust for multiple testing, we applied a Bonferroni correction based on the number of pQTLs analyzed.

SMR^43,44^ was conducted to evaluate the potential causal effects of DNA methylation changes on gSEM-derived suicide traits in individuals of EUR ancestry. mQTL data were derived from brain and blood tissues, with the 1000 Genomes Project Phase 3 data of EUR ancestry used as the reference panel. Brain-specific effects were assessed with the mMeta-brain mQTL data,^67^ which integrated data from (1) dorsolateral prefrontal cortex (ROSMAP),^68^ (2) nondissected fetal brain,^69^ and (3) frontal cortex of adult brain.^70^ To further assess fetal-specific effects, SMR was also performed using the fetal brain mQTL data alone. Blood mQTL resources included the GoDMC catalog,^71^ the ARIES project,^72^ and datasets from Hatton et al.,^73^ McRae et al.,^74^ and Hannon et al.^75,76^ SMR analysis targeted cis-mQTLs within ±2Mb of each cytosine-phosphate-guanine (CpG) site, with other parameters set to default. To improve statistical power, we performed a sample-size-weighted meta-analysis using METAL^34^ to combine results from all brain and blood datasets, including genomic control and Z-statistic-based sample overlap correction. Multiple testing correction was applied using FDR approach to account for the number of CpG sites tested, and significant methylation signals were defined as FDR q<0.05 and HEIDI heterogeneity test *P*>0.05, which were then annotated to the corresponding genes.

Integrating evidence from all five gene-discovery analyses described above, we prioritized genes associated with suicide traits. A gene was assigned a score of 1 if it reached significance in any tissue- or dataset-specific or combined analysis in one approach. Prioritization was conducted separately for SI, SA, and SP, and then the scores were summed across the three traits. To further validate the prioritized genes through both regulatory and coding evidence, we performed genomic annotation for EUR combined suicide GWAS signals using the VEP tool.^61^

A gene-set enrichment analysis was further performed using GSA-MiXeR^45^ to investigate the gene functional annotation associated with gSEM derived suicide traits in EUR ancestry. This approach estimates the fold enrichment of heritability across 79 253 predefined GO terms, while accounting for LD structure using the 1000 Genomes Project Phase 3 EUR reference panel.^58^ The tested GO terms included three categories: biological processes (BP), molecular functions (MF), and cellular components (CC). Bonferroni correction was applied, considering the total number of GO terms evaluated. For GO terms achieving Bonferroni significance, we further calculated the heritability-fold-enrichment differences between pairs of the three suicide traits, with FDR q_diff_<0.05 considered as significant. To identify representative functional themes, we reduced redundancy among significant GO terms with fold enrichment>1 within each GO category using rrvgo^77^, considering a similarity threshold of 0.7. rrvgo selected the representative term within each similarity group as the one with the highest score, defined as the minus log-transformed *P*-value.

We used a genetically informed drug repurposing framework, DRUGSETS,^46^ to identify potential therapeutic candidates for suicide traits. Gene targets and drug–gene interactions were obtained from the Drug Gene Interaction Database and the Clue Repurposing Hub. Drug-gene sets were created for 1201 drugs. Competitive gene-set analysis was conducted with MAGMA v1.10,^38^ conditioning on a background set of 2281 drug target genes, to test for significant associations between drug–gene sets and suicide traits. Bonferroni correction was applied across the 993 drug–gene sets tested to account for multiple testing. In subsequent analyses, drug–gene sets were grouped by Anatomical Therapeutic Classification (ATC) III code, clinical indication, and mechanism of action (MOA). Multiple linear regression models were used to test whether membership in these drug groups predicted stronger associations, with drug group membership as the predictor and MAGMA t-statistics from drug–gene set analyses as the outcome.

### Phenome-wide analyses

We first estimated genetic correlations between gSEM derived suicide traits and six psychiatric disorders in EUR ancestry using LDSC.^47^ These included major depressive disorder (MDD),^78^ post-traumatic stress disorder (PTSD),^79^ anxiety (ANX),^80^ bipolar disorder (BIP),^81^ schizophrenia (SCZ),^82^ and substance use disorders (SUD).^83^ In addition, we investigated the shared variants between suicide traits and these psychiatric disorders by applying bivariate MiXeR^49^. We used the 1000 Genomes Project Phase 3 data of EUR ancestry as the reference panel,^58^ and retained only those results with best_vs_min_AIC>0 and/or best_vs_max_AIC>0.

To account for the potential effects of these psychiatric disorders on suicide GWAS, we subsequently performed a conditional analysis using mtCOJO,^84^ a multitrait-based conditional and joint analysis method implemented in the GCTA software package.^60^ Briefly, mtCOJO estimated the SNP effects on suicide traits while conditioning on each psychiatric disorder, and this approach is robust to sample overlap. Genome-wide significant and LD-independent SNPs (*P*<5×10^-8^, r^2^<0.1) from each psychiatric disorder GWAS were used as instruments to estimate the conditional effects on the suicide traits. Following mtCOJO analysis, we re-estimated the SNP-based heritability of gSEM derived suicide traits in EUR ancestry, assessing their genome-wide associated statistics conditioned for each psychiatric disorder through LDSC^22^.

We next extended the LDSC analysis to a phenome-wide scale by estimating genetic correlations of suicide traits with 11 374 complex traits from three large cohort resources: Pan-UKB (7153 traits),^54^ FinnGen (Release 12; 2469 traits),^30^ and MVP (1752 traits).^85^ In addition, we assessed genetic correlations with 233 metabolomic traits meta-analyzed from UKB^86^ and non-UKB^87^ GWAS datasets using METAL, as well as with 3935 brain imaging-derived phenotypes (IDP) from UKB.^88^ Beyond the primary analysis, we re-evaluated these genetic correlations using GWAS summary statistics for suicide traits after conditioning psychiatric disorders via mtCOJO. Bonferroni correction was applied to account for the number of complex traits (N_trait_=11 374) in three cohorts and the number of metabolomic traits (N_trait_=233) separately. Given the relatively small sample sizes of brain IDP GWASs (up to 33 224 participants), a less stringent significance threshold (*P*<0.01) was used. Across all analyses, only traits with robust SNP-based heritability (SNP-h^2^ Z>4) were retained in the results.

For traits showing significant genetic correlations in the primary LDSC analysis, we further assessed their potential bidirectional causal relationships with suicide traits using three complementary approaches: LCV,^50^ MRlap,^51^ and GSMR2.^52^ LCV estimated the genetic causality proportion (GCP) and genetic correlations (ρ) while accounting for sample overlap between the two traits. LD scores were pre-computed from the 1000 Genomes Project phase 3 EUR data. A negative GCP indicates that the suicide trait is the outcome of another complex trait, whereas a positive GCP indicates the reverse direction. We applied a nominal significance threshold (*P*<0.05), as LCV is more stringent and conservative than MR.^50^ MRlap provides a corrected causal effect estimate by simultaneously accounting for potential sample overlap, weak instrument bias, and winner’s curse. When selecting instrumental variables (IVs), we used a *P*-value threshold of 5×10^-8^ for suicide traits and 1×10^-6^ for other complex traits, given the relatively smaller sample sizes. Other parameters were set to default. In GSMR2, we excluded cohort-specific suicide GWAS data having sample overlap with other complex traits and re-conducted gSEM (≥3 GWASs) or METAL (<3 GWASs) analyses. The same LD reference panel as GCTA-COJO derived from UKB EUR genotype data (N=5000) was used. For IV selection, we applied a *P*-value threshold of 5×10^-7^ for suicide traits and 1×10^-6^ (restricted to the top 1500 variants) for other complex traits. Other parameters were set as follows: multi_snps_heidi_thresh=0.01, nsnps_thresh=5, ld_r2_thresh=0.05, ld_fdr_thresh=0.05, and gsmr2_beta=1. In both MRlap and GSMR2, Bonferroni correction was applied to account for multiple tests (N=6670) for complex traits in Pan-UKB, FinnGen, and MVP data, while *P*<0.01 was used to identify significant metabolomic traits and brain IDPs.

## Results

### Genome-wide significant loci

Across the three cohorts of SI in EUR ancestry, UKB showed the highest SNP-based heritability on the liability scale (SNP-h^2^=0.072±0.005), while among the four cohorts of SA in EUR ancestry, FinnGen had the highest heritability (SNP-h^2^=0.140±0.009) (appendix pp 1 Table S2A). Genetic correlations among these seven GWASs in EUR ancestry ranged from 0.238 to 0.862, all significant after Bonferroni multiple testing correction (*P*<0.002; appendix pp 3 Figure S1; appendix pp 1 Table S2B). Consequently, we applied gSEM to combine the SI and SA GWASs in EUR ancestry separately, as well as to integrate all seven EUR GWASs into a new phenotype SP. Parallel analysis suggested a one-factor model across SI, SA, and SP in gSEM analyses (appendix pp 4–5 Figure S2–3). The gSEM-SA factor showed higher heritability (SNP-h^2^=0.115±0.005) compared to the gSEM-SI (SNP-h^2^=0.040±0.002) and gSEM-SP factors (SNP-h^2^=0.050±0.002), and SA–SP genetic correlation (r_g_=0.960, *P*<1×10^-300^) was the highest, followed by SI–SP (r_g_=0.865, *P*<1×10^-300^) and SI–SA (r_g_=0.639, *P*=6.81×10^-135^). Using METAL, we performed meta-analysis for each non-EUR ancestry and across-ancestry meta-analysis for SI and SA. However, the heritability estimates of meta-analyzed GWASs were not statistically significant (*P*>0.05) in non-EUR ancestries.

In total, we identified 87 independent lead SNPs (*P*<5×10^-8^) from 80 loci considering all three suicide traits, of which 49 SNPs were novel (Figure 1; appendix pp 1 Table S3A). Most identified SNPs were intronic (36 SNPs) or intergenic (31 SNPs). The most significant novel lead SNP was rs2093599 (*P*=2.52×10^-11^; intergenic; nearest gene: *LINC01065*), followed by rs1008151 (*P*=3.35×10^-11^; intronic; nearest gene: *METAP1D*) and rs4425103 (*P*=1.48×10^-10^; intergenic; nearest gene: *AC096570.1*). These loci have previously been reported to be associated with multiple traits, such as insomnia, educational attainment, and MDD (appendix pp 1 Table S3B). Regarding each ancestry, genome-wide significant SI and SA lead SNPs were identified in EUR (SI: 12 SNPs; SA: 31 SNPs), AFR (SI: 2; SA: 1), AMR (SI: 2; SA: 3), and ASN/EAS (SI: 1; SA: 1) (appendix pp 1 Table S3C–D). In SI cross-ancestry meta-analysis, 15 lead SNPs in 14 loci were identified from 283 genome-wide significant SNPs (*P*<5×10^-8^; 94 SNPs at *P*<5×10^-9^), with five additional lead SNPs identified exclusively in EUR (2 SNPs), AMR (2), and ASN (1) ancestries (appendix pp 6 Figure S4; appendix pp 1 Table S3E). The most significant lead SNP was rs62474669 (*P*=1.23×10^-11^; intergenic; nearest gene: *AC068610.5*). In SA cross-ancestry meta-analysis, 32 lead SNPs in 29 loci were identified from 818 significant SNPs (*P*<5×10^-8^; 451 SNPs at *P*<5×10^-9^), with five additional lead SNPs identified exclusively in EUR (4 SNPs) and AMR (1) (appendix pp 6 Figure S4; appendix pp 1 Table S3F). The most significant lead SNP was rs12666306 (*P*=1.26×10^-16^; intergenic; nearest gene: *RP11-222O23.1*). For gSEM-SP factor in EUR, 61 independent SNPs in 54 loci were recognized from 1921 significant SNPs (*P*<5×10^-8^; 982 SNPs at *P*<5×10^-9^), with the most significant lead SNP being rs62474669 (*P*=1.16×10^-20^; appendix pp 1 Table S3A).

**Figure 1:**
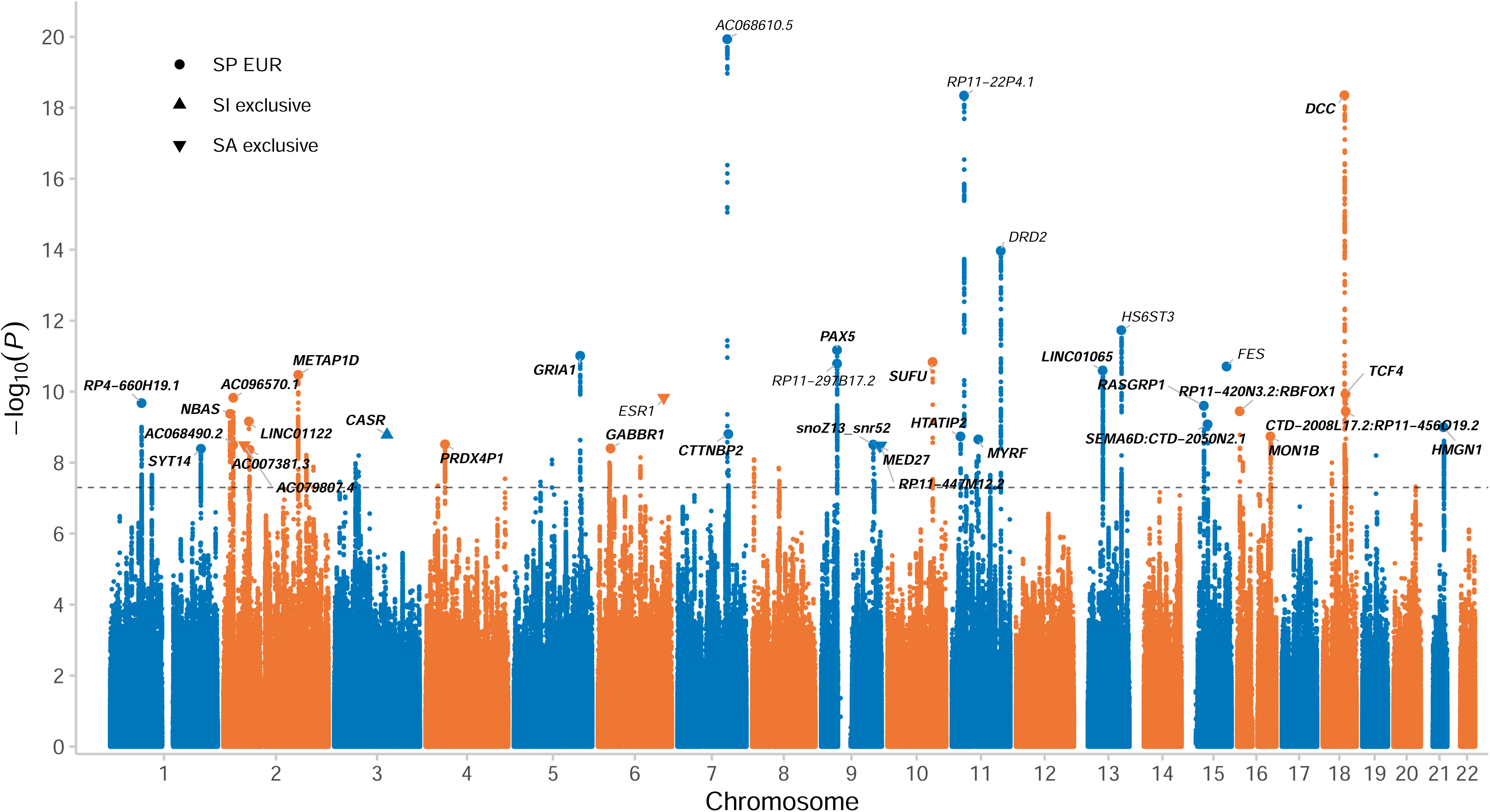
Manhattan plot of genome-wide association studies for suicide phenotypes. Manhattan plot is shown for genome-wide association study (GWAS) of suicidality spectrum (SP) in individuals of European ancestry, with additional independent lead SNPs identified for suicide ideation (SI) and suicide attempt (SA) across ancestries. Lead SNPs with *P*<5×10^-9^ are highlighted and mapped to the nearest genes; gene symbols in bold indicate novel loci not previously reported. The horizontal dashed line represents the genome-wide significance threshold (*P*=5×10^-8^).

GCTA-COJO joint analysis identified two additional SNPs for SA (rs7229145, *P*_j_=2.09×10^-8^, intergenic; rs10853530, *P*_j_=1.4×10^-8^, intronic, mapped gene: ENSG00000267101) and one additional SNP for SP (rs3909543, *P*_j_=3.59×10^-8^, intronic, mapped gene: *PMFBP1*) in EUR ancestry (appendix pp 1 Table S4), which were independent (LD r^2^<0.1) with the lead SNPs identified by FUMA. However, these additional SNPs were not independent of previously reported loci.

In fine-mapping analysis, SuSiEx identified six putative cross-ancestry causal SNPs (PIP>0.8; three SNPs with PIP>0.95) within 16 95%-credible sets for SI, seven putative causal SNPs (PIP>0.8; five SNPs with PIP>0.95) within 31 95%-credible sets for SA, and three putative causal SNPs (PIP>0.8; two SNPs with PIP>0.95) within 47 95%-credible sets for SP. Notably, four SNPs showed PIP values approaching 1: rs74815327 (mapped to *LINC02790*), rs145491971 (*GOLGA4*), rs7461966 (ENSG00000253500), and rs7177338 (*FES*) (Table 1; appendix pp 1 Table S5).

**Table 1:** Credible sets and putative causal SNPs for suicide traits identified by multi-ancestry fine-mapping.

| Suicide phenotype | CHR | CS No. | CS purity | Max PIP | Max PIP SNP | BP | Variant function | Mapped gene symbol ID | Ensembl gene ID | Gene type |
| --- | --- | --- | --- | --- | --- | --- | --- | --- | --- | --- |
| Suicide ideation | 1 | 1 | 1.000 | 1.000 | rs74815327 | 96452462 | Intron | *LINC02790* | ENSG00000237435 | lncRNA |
|  | 2 | 2 | 0.941 | 0.847 | rs34552237 | 103438197 | Downstream gene | *TMEM182* | ENSG00000170417 | Protein coding |
|  | 2 | 3 | 0.969 | 0.813 | rs112432457 | 172901195 | Intron | *METAP1D* | ENSG00000172878 | Protein coding |
|  | 3 | 4 | 0.941 | 0.942 | rs145321328 | 121926006 | Intron | *CASR* | ENSG00000036828 | Protein coding |
|  | 6 | 5 | 1.000 | 0.964 | rs145624415 | 116080621 | Intergenic | - | - | - |
|  | 9 | 6 | 1.000 | 0.969 | rs9407059 | 37021165 | Intron | *PAX5* | ENSG00000196092 | Protein coding |
| Suicide attempt | 2 | 1 | 1.000 | 0.996 | rs73065164 | 210290180 | Intron | *MAP2* | ENSG00000078018 | Protein coding |
|  | 2 | 2 | 0.989 | 0.905 | rs12473936 | 48273872 | Intron | - | ENSG00000230773 | lncRNA |
|  | 3 | 3 | 1.000 | 1.000 | rs145491971 | 37293763 | Intron | *GOLGA4* | ENSG00000144674 | Protein coding |
|  | 8 | 4 | 1.000 | 1.000 | rs7461966 | 88741064 | Intron | - | ENSG00000253500 | lncRNA |
|  | 9 | 5 | 0.917 | 0.896 | rs144908548 | 25888160 | Downstream gene | - | ENSG00000300408 | lncRNA |
|  | 14 | 6 | 1.000 | 0.988 | rs860324 | 57348922 | Intron | *OTX2-AS1* | ENSG00000248550 | lncRNA |
|  | 15 | 7 | 1.000 | 1.000 | rs7177338 | 91428636 | Intron | *FES* | ENSG00000182511 | Protein coding |
| Suicidality spectrum | 15 | 1 | 1.000 | 0.991 | rs7177338 | 91428636 | Intron | *FES* | ENSG00000182511 | Protein coding |
|  | 18 | 2 | 1.000 | 0.988 | rs2852978 | 53749583 | Intron | *LINC01539* | ENSG00000267712 | lncRNA |
|  | 19 | 3 | 0.967 | 0.894 | rs8105753 | 31927547 | Intergenic | - | - | - |

Fine-mapping was performed using SuSiEx. SNPs with PIP > 0.8 were considered putative causal variants. CHR=chromosome.
CS=credible set. PIP=posterior inclusion probability. SNP=single-nucleotide polymorphism. BP=base pair position.

### Gene prioritization

In MAGMA analysis, we observed that SI, SA, and SP GWASs were enriched for multiple tissue-specific transcriptomic regulations after Bonferroni correction (*P*<9.43×10^-4^; appendix pp 7 Figure S5; appendix pp 1 Table S6). Except for EBV-transformed lymphocytes for SA in the cross-ancestry meta-analysis, all enriched tissues were brain-related, with the most significant enrichment observed in cerebellum for all three suicide traits (EUR SI, cerebellar hemisphere *P*=4.01×10^-5^; cross-ancestry SA, cerebellar hemisphere *P*=1.57×10^-6^; EUR SP, cerebellum *P*=6×10^-8^). However, in EUR SA GWAS, the most significant enrichment was in frontal cortex BA9 (*P*=7.55×10^-6^).

MAGMA gene-based analysis identified 24 genes for SI (EUR: 22 genes, *P*<2.74×10^-6^; AMR: 1 gene, *P*<2.65×10^-6^; cross ancestry: 2 genes, *P*<2.64×10^-6^), 46 genes for SA (EUR: 40 genes, *P*<2.73×10^-6^; AFR: 6 genes, *P*<2.68×10^-6^; AMR: 1 gene, *P*<2.71×10^-6^; cross ancestry: 14 genes, *P*<2.67×10^-6^), and 85 genes for SP (EUR: *P*<2.74×10^-6^) after Bonferroni correction (appendix pp 8 Figure S6; appendix pp 1 Table S7). For all three suicide traits, the most significant gene was *DCC* (*P*_SI_=1.02×10^-13^, *P*_SA_=2.44×10^-13^, *P*_SP_=2.46×10^-22^).

The brain-specific cross-tissue TWAS identified 7 genes for SI (most significant: *NR1H3*, *P*_brain_=7.19×10^-10^), 7 genes for SA (most significant: *DNAJC3*, *P*_brain_=4.44×10^-8^), and 32 genes for SP (most significant: *RP11-24H2.3*, *P*_brain_=4.32×10^-11^) after Bonferroni correction (*P*<2.32×10^-6^; appendix pp 9–12 Figure S7–10; appendix pp 1 Table S8A–C). When 36 non-brain body tissues were also included, 10, 23, and 77 additional genes were identified for SI, SA, and SP, respectively (appendix pp 9–12 Figure S7–10; appendix pp 1 Table S8A–C). Among these, *RMC1* (synonym: *C18orf8*) for SI (significant in 3 brain and 5 body tissues), and *GMPPB* for both SA (in 10 brain and 25 body tissues) and SP (in 12 brain and 25 body tissues) showed the highest number of tissue-specific associations. Regarding brain tissue-specific analysis, the top tissues with the most associations for SI were frontal cortex BA9 (5 genes), cerebellar hemisphere (4 genes), and cerebellum (4 genes). For SA, the top tissues were cerebellum (8 genes), cerebellar hemisphere (6 genes), and putamen of basal ganglia (4 genes). For SP, cerebellum (15 genes), caudate of basal ganglia (12 genes), and cerebellar hemisphere (11 genes) showed the greatest number of associations.

isoTWAS identified 46, 71, and 117 FDR-significant genes for SI, SA, and SP, respectively (appendix pp 13 Figure S11; appendix pp 1 Table S9A–C). For both SI and SP, the most significant gene was *CLYBL* (SI screening *P*=9.34×10^-68^; SP screening *P*=3.74×10^-127^). For SA, *ZNF530* (Screening *P*=5.6×10^-180^) was the most significant gene, with *CLYBL* also ranking as the second most significant gene (Screening *P*=6.68×10^-62^).

PWAS identified 42 Bonferroni-significant proteins in total: 17 for SI (*P*<2.81×10^-5^), 14 for SA (*P*<2.77×10^-5^), and 33 for SP (*P*<2.8×10^-5^) (Figure 2; appendix pp 14 Figure S12; appendix pp 1 Table S10). SLC25A12 was the most significant protein for both SI (Z_meta_=6.892, *P*=5.49×10^-12^) and SP (Z_meta_=8.367, *P*=5.9×10^-17^), while GMPPB was the most significant for SA (Z_meta_=8.023, *P*=1.03×10^-15^). There were 12 proteins showing consistent associations between SI and SP, and 10 proteins between SA and SP. However, comparing SI and SA, most proteins were exclusively associated with either phenotype, with CTNND1 being the only protein significantly associated with both SI and SA.

**Figure 2:**
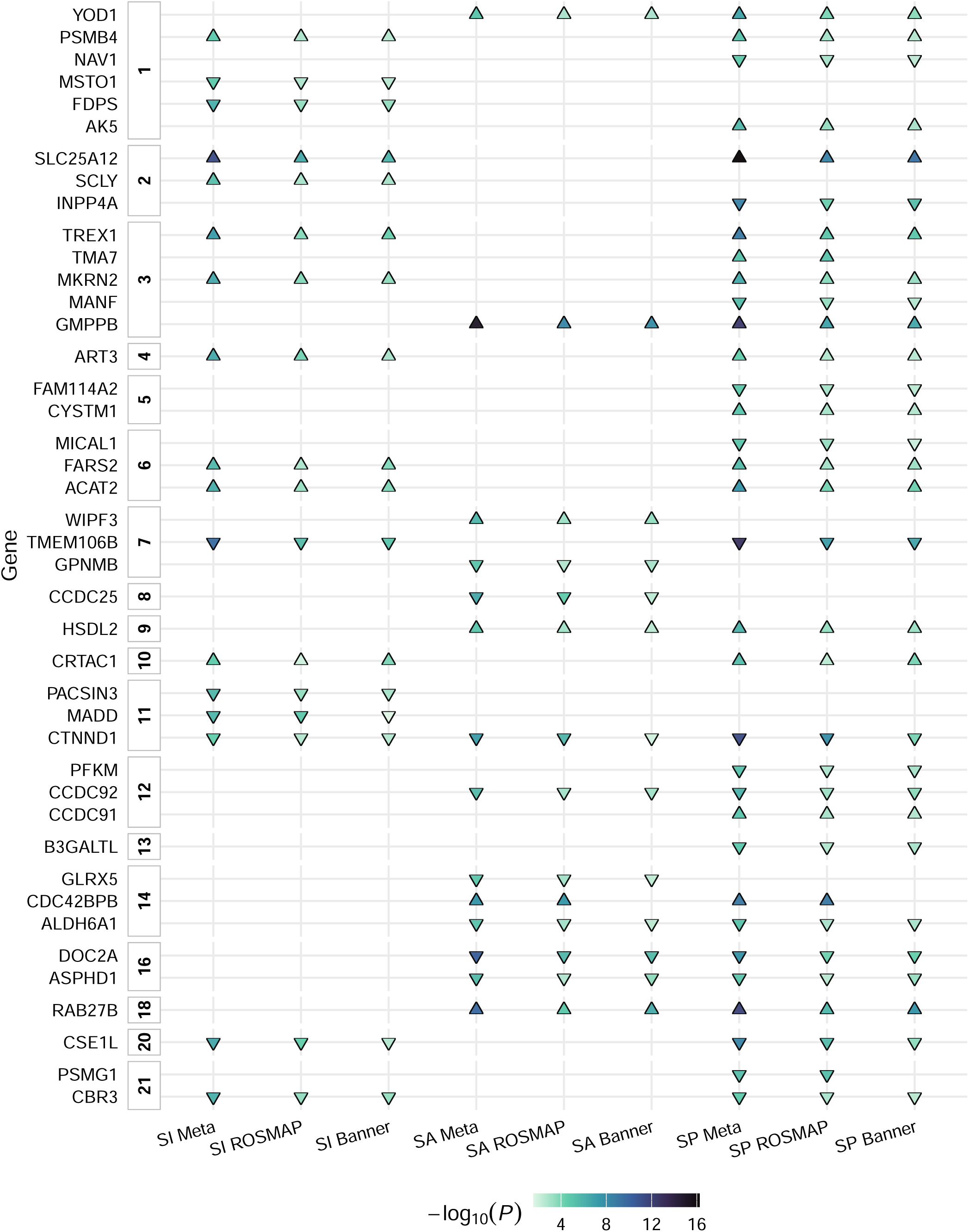
Significant genes identified in proteome-wide association studies for suicide phenotypes. The plot shows significant genes identified in meta-analysis of proteome-wide association studies (PWAS) based on the Religious Orders Study and Memory and Aging Project (ROSMAP) and Banner Sun Health Research Institute (Banner) datasets. Upward and downward triangles represent positive and negative z-scores, respectively. SI=suicide ideation. SA=suicide attempt. SP=suicidality spectrum.

SMR meta-analysis integrating brain and blood mQTL datasets identified 23 significant CpG sites for SI (most significant: cg13810695, *P*_meta_=2.3×10^-12^), 114 CpGs for SA (most significant: cg04339947, *P*_meta_=4.02×10^-16^), and 277 CpGs for SP (most significant: cg20866694, *P*_meta_=1.37×10^-16^) after FDR multiple testing correction (appendix pp 15 Figure S13; appendix pp 1–2 Table S11A–C). Fetal-specific signals were predominantly observed for SP, with the strongest association being at cg13550731 (gene annotation: *DYNC1I2*). In dataset-specific analysis, 211, 690, and 2163 CpG sites were significant in at least one dataset for SI, SA, and SP, respectively. The CpG sites with the highest number of significant datasets were cg20866694 (significant in 6 datasets) for SI, cg11890956 (in 7 datasets; gene annotation: *PSMG1*) for SA, and cg12091567 (in 7 datasets; gene annotation: *LOC651250*) for SP. Among these signals, multiple CpG sites were annotated to the same genes, such as *LOC645323* (10 sites) and *RERE* (7 sites) for SI, *MAD1L1* (29 sites) and *C6orf41* (11 sites) for SA, and *MAD1L1* (44 sites) and *TRIM26* (28 sites) for SP.

Integrating gene-level evidence, we identified a total of 1052 genes, with the top three genes (*UGGT2*, *GMPPB*, and *BRWD1*) significant in eight analyses (Figure 3; appendix pp 16 Figure S14; appendix pp 2 Table S12). Among these, 16 genes had evidence supporting associations with all three suicide traits (*UGGT2*, *ANKK1*, *METAP1D*, *DCC*, *CTNND1*, *ALG12*, *PAX5*, *GRIA1*, *STYXL1*, *RBFOX1*, *CLYBL*, *KTN1-AS1*, *DNAJC24*, *SFXN2*, *ARL3*, *MDH2*). A total of 65, 114, and 537 genes were exclusively associated with SI (e.g., *NUP160*, *H2BC5*/*HIST1H2BD*, *PTPMT1*, *GJC1*, *FDPS*), SA (e.g., *ALDOA*, *PEX7*, *PPP4C*, *RP1-45I4.3*, *MAP3K5*), or SP (e.g., *SMPD2*, *MICAL1*, *METTL21A*, *TMA7*, *H2BC15*/*HIST1H2BN*), respectively, with evidence from at least one analysis. For SP GWAS signals, the VEP tool identified two missense variants located in *ANKK1* (rs2734849) and *SYCE1L* (rs62049594), with the two genes having integrated evidence scores of 7 and 4, respectively. We also identified four synonymous variants in three genes: *MON1B* (SA and SP: rs2232504), *PSMG1* (SP: rs14194 and rs3171465), *DRD2* (SP: rs6277), with evidence scores≥6.

**Figure 3:**
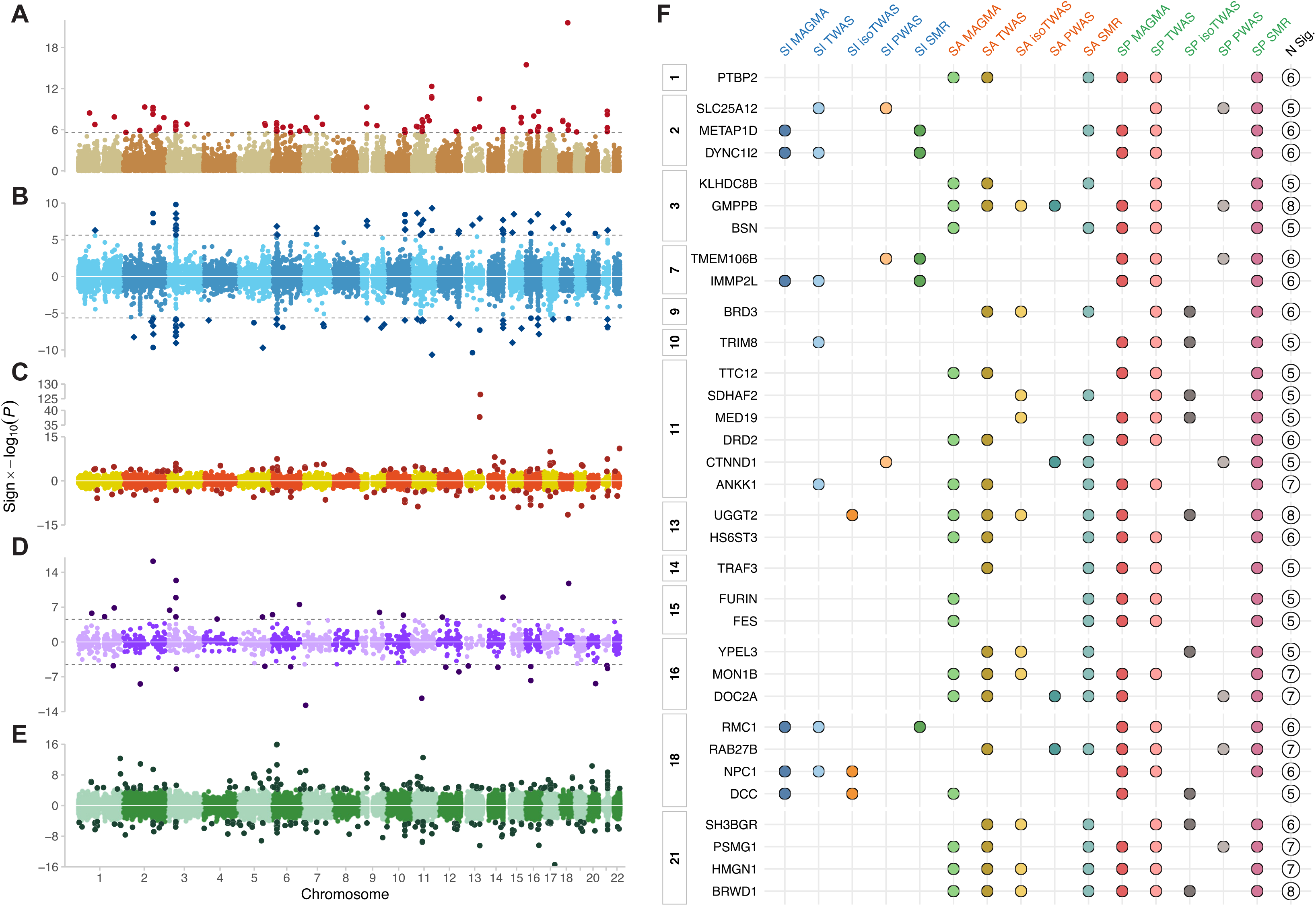
Prioritized Genes integrating evidence from five gene-discovery analyses across suicide phenotypes. A: MAGMA gene-based analysis; B: transcriptome-wide association study (TWAS). Circles represent genes from multi-tissue analysis using 13 GTEx brain tissues, while diamonds indicate additional significant genes identified when all 49 GTEx tissues were included; C: isoform-level TWAS; D: proteome-wide association study (PWAS); E: methylation quantitative trait loci-based summary-data Mendelian randomization (mQTL-SMR); F: gene prioritization scores. Panels A–E show results of the five gene-discovery approaches for suicidality spectrum. In Panels B–E, y-axis values represent -log_10_(*P*), with positive and negative values corresponding to the sign of the Z-value. Panel F lists genes with a prioritization score≥5. For each approach and for each suicide trait, a gene was assigned one point if it showed a significant association in any tissue, dataset, or multi-analysis, and the points were then summed across the three suicide traits and five approaches. SI=suicide ideation. SA=suicide attempt. SP=suicidality spectrum.

### Gene-set enrichment and potential drug candidates

In GSA-MiXeR analysis, SNP-based heritability was significantly enriched in 1121, 1346, and 1595 GO terms for SI, SA, and SP, respectively, after Bonferroni correction (*P*<6.31×10^-7^; appendix pp 2 Table S13A). Comparing SI and SA, apoptotic process involved in development showed a significant difference in fold enrichment after FDR correction (SI fold enrichment=0.211, SA fold enrichment=1.268, *P*_diff_=2.4×10^-5^). Using rrvgo, we clustered those significant terms with positive fold enrichment into 95 non-redundant groups for SI, 103 for SA, and 104 for SP (appendix pp 17 Figure S15; appendix pp 2 Table S13B). Several high-scored groups were identified across suicide traits, such as cellular response to stress, Golgi apparatus, and transcription regulator activity.

DRUGSETS analysis suggested four potential therapeutic drug candidates for SA after Bonferroni correction (*P*_drug-set_<5.04×10^-5^): cariprazine (*P*_drug-set_=2.93×10^-5^), droperidol (*P*_drug-set_=1.22×10^-5^), molindone (*P*_drug-set_=4.29×10^-5^), and paliperidone (*P*_drug-set_=8.26×10^-6^), as well as two candidates for SP: cariprazine (*P*_drug-set_=4.55×10^-5^) and chlorprothixene (*P*_drug-set_=4.81×10^-5^) (appendix pp 2 Table S14). MOA enrichment analysis revealed that the four SA candidate drugs were significantly enriched for topoisomerase inhibitor (*P*_enrichment_=1.12×10^-4^), while the SP candidate drug was enriched for sodium channel blocker (*P*_enrichment_=2.67×10^-5^). No significant drug candidate was identified for SI.

### Suicide-related pleiotropy

We investigated genetic correlations of SI, SA, and SP with six psychiatric disorders and found that all pairs were significant after Bonferroni correction (*P*<8.3×10^-3^; Figure 4A; appendix pp 2 Table S15). For SI, MDD showed the highest genetic correlation (r_g_=0.69, *P*=4.59×10^-266^), while for SA and SP, MDD (SA r_g_=0.793, *P*<1×10^-300^; SP r_g_=0.846, *P*<1×10^-300^), PTSD (SA r_g_=0.777, *P*<1×10^-300^; SP r_g_=0.791, *P*<1×10^-300^), and ANX (SA r_g_=0.789, *P*=5.45×10^-302^; SP r_g_=0.781, *P*<1×10^-300^) had a higher genetic correlation compared to BIP, SCZ, and SUD. When comparing the suicide traits, SA and SP consistently showed higher genetic correlations than SI across all six psychiatric disorders. Although SP showed a higher genetic correlation with MDD than SA, the opposite was observed for SUD.

**Figure 4:**
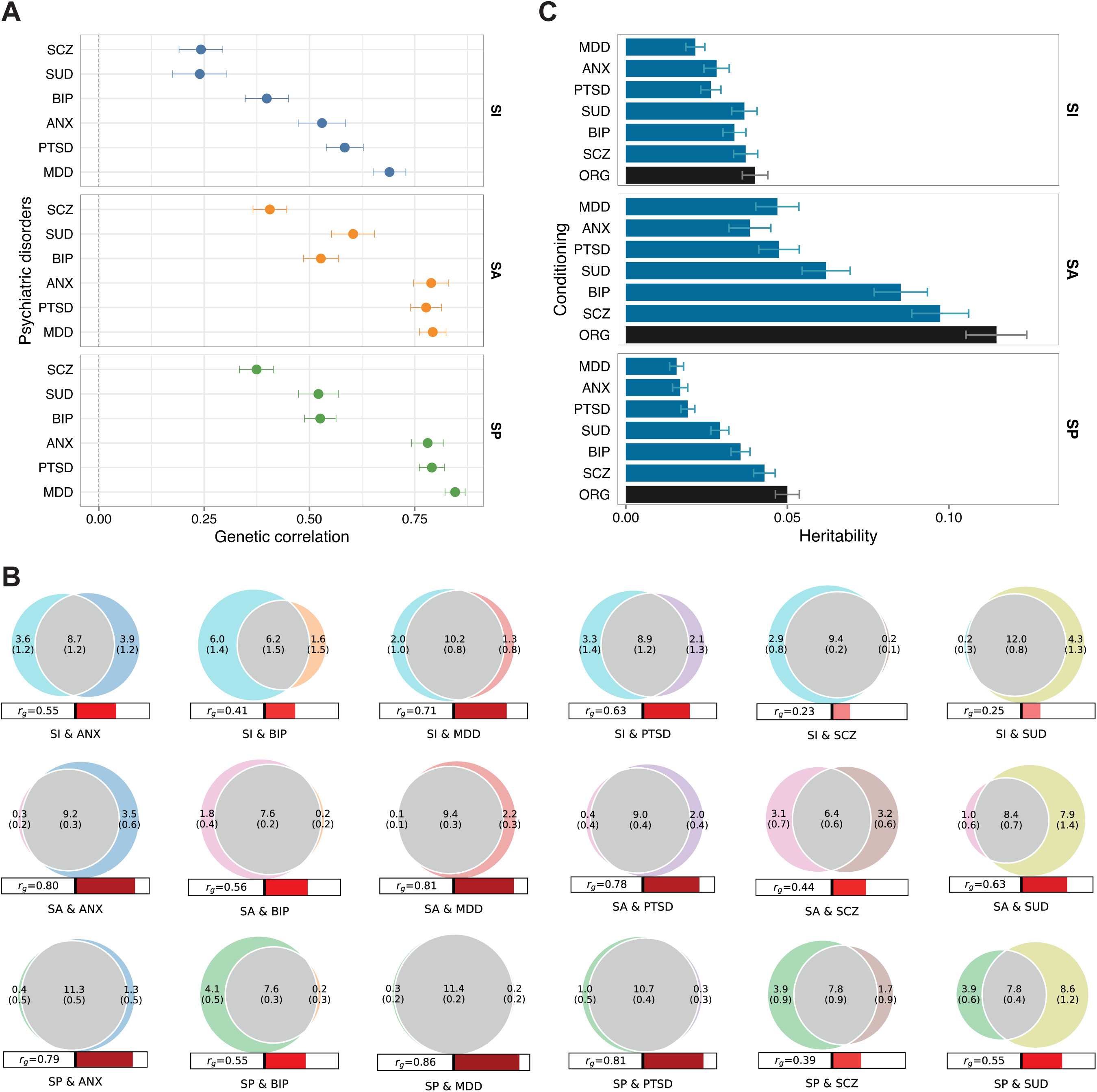
Genetic relationships between suicide phenotypes and psychiatric disorders. A: Genetic correlations. B: SNP-based heritability of suicide phenotypes in individuals of European ancestry before and after conditioning on each psychiatric disorder. C: Venn plots of variants shared between suicide phenotypes and psychiatric disorders. SI=suicide ideation. SA=suicide attempt. SP=suicidality spectrum. ORG=original heritability without conditioning. MDD=major depressive disorder. PTSD=post-traumatic stress disorder. ANX=anxiety. BIP=bipolar disorder. SCZ=schizophrenia. SUD=substance use disorders.

Based on MiXeR bivariate modeling, MDD showed the largest proportion of shared causal variants with all three suicide traits (MDD–SI: 86%, MDD–SA: 89%, MDD–SP: 98%; Figure 4B; appendix pp 18 Figure S16; appendix pp 2 Table S16) when considering all variants jointly between each pair. The fraction of concordant effects among the shared variants was greater than 80% for all MDD-related pairs (MDD–SI: 82%, MDD–SA: 86%, MDD–SP: 84%). Notably, although MiXeR indicated that SCZ and SUD had relatively low genetic correlations with SI (SCZ–SI r_g_: 0.234±0.005, SUD–SI r_g_: 0.254±0.016), both disorders showed a relatively high proportion of shared variants with SI (SCZ–SI proportion: 86%, SUD–SI proportion: 84%). A similar pattern was observed for BIP and SA (r_g_: 0.56; proportion: 88%). The highest fractions concordant within shared variants were found for SUD–SP (94%) and SUD–SA (91%), despite their relatively low proportions of shared variants (SUD–SP: 56%, SUD–SA: 65%).

In phenome-wide genetic correlation analysis, we investigated the relationships between the three suicide traits and 11 374 complex traits from UKB, FinnGen, and MVP. The analysis identified 449, 1497, and 1389 traits significantly correlated with SI, SA, and SP, respectively, after Bonferroni correction (*P*<4.4×10^-6^; appendix pp 19 Figure S17; appendix pp 2 Table S17). Most significant correlations involved mental health traits, such as depression, anxiety, BIP, SUD, personality disorders, and psychosocial adversity. In addition, we also observed significant genetic correlations between suicide traits and a wide range of physical health traits (e.g., pain/musculoskeletal, cardiometabolic, gastrointestinal, respiratory/allergic, urologic/renal, sleep/fatigue, neurological/cognitive, endocrine, gynecological, dental, otolaryngologic/ocular, infectious conditions, and biomarkers), behaviors (e.g., tobacco smoking, alcohol drinking, cannabis use, sleep, mobile/computer/TV use, physical activity, diet habits, commuting pattern, occupational/sun exposure, and sexual behaviors), and socioeconomic factors (e.g., Townsend deprivation index, low education/income levels, unemployment, lack of social support/relationships, household/housing characteristics, and family history). Beyond traits from the three comprehensive cohorts, we also estimated genetic correlations of suicide traits with 233 metabolomic traits and 3935 brain IDPs. The results showed 50 and 44 metabolomic traits correlated with SA and SP (*P*<2.15×10^-4^), and 19, 163, and 48 brain IDPs correlated with SI, SA, and SP, respectively (*P*<0.01). The top five metabolomic traits with the most significant genetic correlations included: phospholipids to total lipids ratio in large HDL (SA r_g_=0.166, *P*=1.65×10^-10^), ratio of polyunsaturated fatty acids to total fatty acids (SA r_g_=-0.158, *P*=3.24×10^-10^), ratio of monounsaturated fatty acids to total fatty acids (SA r_g_=0.146, *P*=3.71×10^-9^), ratio of polyunsaturated fatty acids to total fatty acids (SP r_g_=-0.132, *P*=5.73×10^-9^), and phospholipids to total lipids ratio in large HDL (SP r_g_=0.144, *P*=1.32×10^-8^). Across brain IDPs, all suicide traits showed convergent genetic correlations with reduced volumes in cerebellar vermis/lobules VIII–IX, right accumbens, hippocampal subfields, lateral occipito-temporal sulcus, and insula, as well as with corticospinal-tract microstructural abnormalities extending into thalamic radiations, long-association fibers, and callosal pathways. The most significant IDPs were weighted-mean L3 in corticospinal tract (left: SA r_g_=0.185, *P*=2.39×10^-5^; right: SA r_g_=0.157, *P*=2.66×10^-5^) and weighted-mean mean diffusivity in corticospinal tract (left: SA r_g_=0.171, *P*=4.45×10^-5^; right: SA r_g_=0.143, *P*=1×10^-4^).

After conditioning on each of the six psychiatric disorders using mtCOJO, the number of significant genetic correlation pairs reduced from 3659 to 585 (conditioned on MDD), 631 (PTSD), 364 (ANX), 3117 (BIP), 3325 (SCZ), and 1464 (SUD) (appendix pp 2 Table S17). For metabolomic traits, no significant correlation remained after conditioning on MDD, PTSD, or ANX. The SNP-based heritability estimates for all suicide traits generally declined after the conditioning but remained significant after Bonferroni correction (*P*<2.78×10^-3^; Figure 4C; appendix pp 2 Table S2A). Conditioning on MDD resulted in the largest decreases for SI (from 0.040±0.002 to 0.022±0.002) and SP (from 0.050±0.002 to 0.016±0.001), while conditioning on ANX showed the largest decrease for SA (from 0.115±0.005 to 0.038±0.003).

To investigate potential causal relationships, we used three complementary approaches (LCV, MRlap, and GSMR2) to assess the genetic correlations identified in the phenome-wide analysis. LCV, MRlap, and GSMR2 identified 211, 439, and 276 traits, respectively, with potential causal effects on suicide traits, as well as 233, 682, and 1088 traits that may be causally affected by suicide traits (LCV: *P*<0.05; MRlap and GSMR2 with UKB/FinnGen/MVP traits: *P*<7.5×10^-6^, with metabolomic traits/brain IDPs: *P*<0.01; appendix pp 2 Table S18–20). Among these, convergent evidence from all three approaches suggested three traits as potential risk factors and 23 traits as potential consequences of suicidal behaviors (Figure 5). Specifically, we identified loneliness (GCP=-0.294±0.128; β_MRlap_=0.297±0.057; β_GSMR2_=0.196±0.036), medical abortion (GCP=-0.214±0.105; β_MRlap_=0.288±0.055; β_GSMR2_=0.163±0.023), and age at first sexual intercourse (GCP=-0.126±0.061; β_MRlap_=-0.092±0.006; β_GSMR2_=-0.454±0.027) as risk factors for SI, SA, and SP, respectively. Regarding potential consequences, multiple drug-use-related mental disorders for SA showed the largest GCP (0.713±0.249; β_MRlap_=0.397±0.086; β_GSMR2_=1.245±0.261). Several traits appeared to be consequences of both SA and SP, including hospital admission (planned, emergency, or without operation), trouble falling asleep, and wheeze or whistling in the chest. Additional consequences primarily included physical conditions (e.g., spinal stenosis, knee internal derangement, essential hypertension), as well as lymphocyte count, anxiety feelings, and father’s chronic bronchitis/emphysema. However, our genetically informed causal inference analyses did not identify any consequences related to SI.

**Figure 5:**
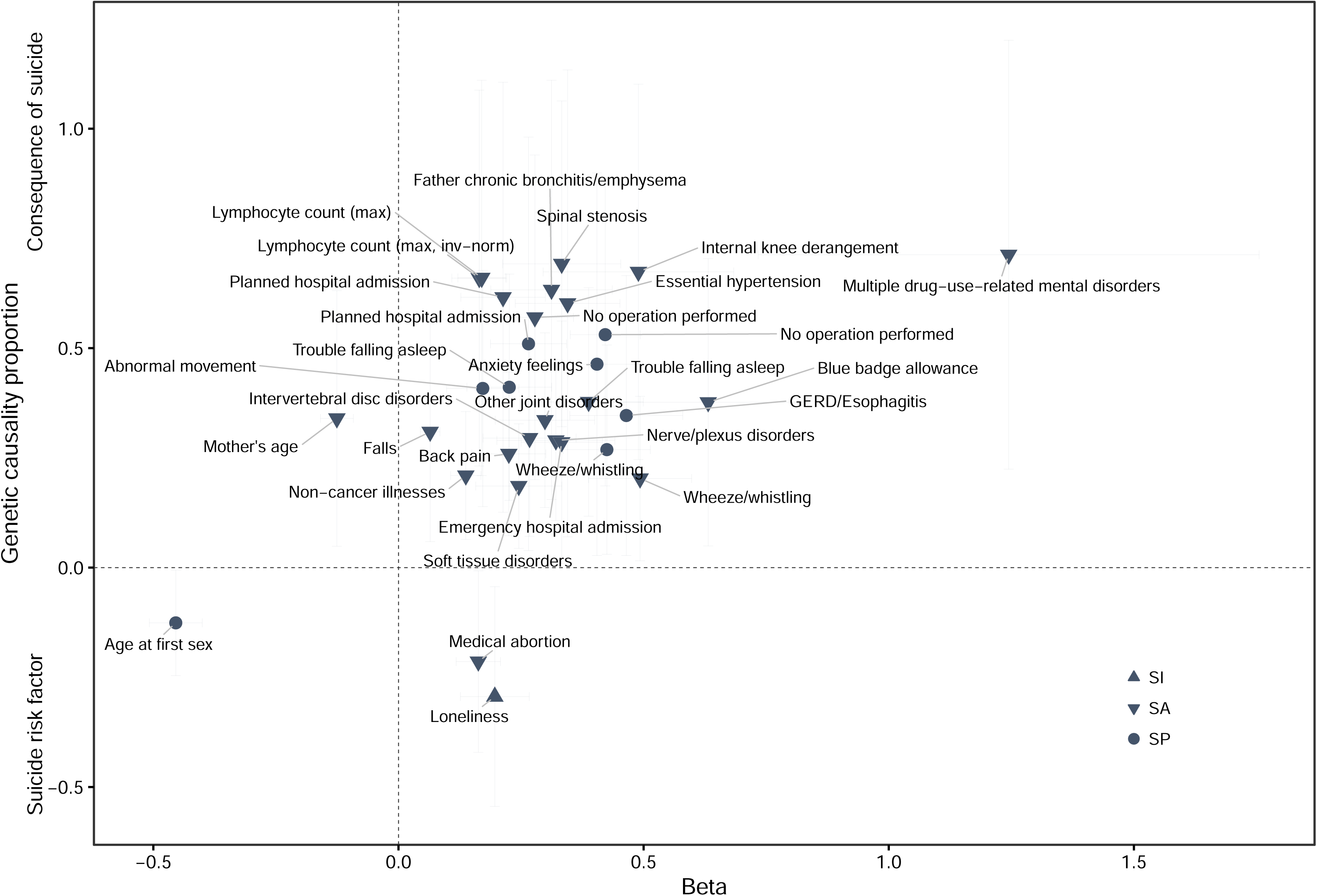
Causal relationships between complex traits and suicide phenotypes. The scatter plot shows complex traits that exhibit significant causal relationships with suicide traits across three complementary methods (LCV, MRlap, and GSMR2). Each point represents a complex trait, mapped by its effect size (beta; x-axis) from GSMR2 and genetic causality proportion (y axis) from LCV. Error bars represent the 95% confidence intervals for both GSMR2 effect sizes and genetic causality proportions. SI=suicide ideation. SA=suicide attempt. SP=suicidality spectrum. GERD: gastroesophageal reflux disease.

## Discussion

In this multi-ancestry GWAS study, we identified 87 independent genomic associations for suicidality, with three additional conditionally independent signals discovered through GCTA-COJO analysis. This represents a substantial increase compared with published studies.^9–19^ When considering the PGC’s most recent GWAS meta-analysis,^20^ 49 of our cross-ancestry associations were novel. Regarding the trait-specific GWAS, SP yielded the largest number of genome-wide associations (61 lead SNPs, compared with 20 for SI and 37 for SA), highlighting the importance of modeling genetic effects shared across suicidal behaviors. In contrast, SNP-based heritability was higher for SA (12%) than for SI (4%) and SP (5%) in EUR. Although SP showed strong genetic correlations with both SI (r_g_=0.87) and SA (r_g_=0.96), we identified 10 SNPs unique to SI and 16 unique to SA beyond the 61 SP-associated SNPs, also indicating the presence of trait-specific effects. Together with the moderate genetic correlation observed between SI and SA (r_g_=0.64), these findings support that their genetic basis is only partially overlapping, as also reported in the recent PGC study.^20^ Besides EUR, we identified genome-wide significant associations for both SI and SA in AFR, AMR, and ASN/EAS populations, greatly expanding the EAS locus identified previously.^20^ However, robust estimation of SNP-based heritability in non-EUR ancestries will require much larger sample sizes.

Among the 16 putative causal SNPs identified through fine-mapping analysis, only rs7177338 was shared in both SA and SP; all other SNPs were trait-specific. For SI, one SNP reached PIP close to 1 and was mapped to a lncRNA gene: *LINC02790*, which has previously been implicated in multiple psychiatric disorders.^89,90^ For SA, two SNPs with PIP close to 1 were mapped to protein-coding genes *GOLGA4* and *FES*. *GOLGA4* is involved in vesicular trafficking at the Golgi apparatus,^91^ and has been associated with gamma-glutamyl transferase levels^92^ and atopic dermatitis,^93^ traits also showed convergent evidence in our phenome-wide genetic correlation analysis of SA and SP. *FES* encodes a tyrosine-specific protein kinase involved in regulating cell differentiation and promoting neurite outgrowth.^94^ It has been associated with SA^17^ and psychiatric disorders, including PTSD,^95^ attention deficit hyperactivity disorder (ADHD),^96^ and schizophrenia.^97^ MAGMA gene expression analysis suggested that cerebellar function may play an important role in suicide onset,^98^ given that cerebellum was the most significant tissue across all suicide traits. Besides brain-derived tissues, we also observed a significant enrichment in EBV-transformed lymphocytes for cross-ancestry SA GWAS, suggesting potential contribution of peripheral immune-related mechanisms.^99^

We applied five complementary approaches to prioritize genes for suicide traits. MAGMA gene-based analysis identified *DCC* as the most significant gene across all suicide traits. *DCC* encodes a netrin 1 receptor involved in axon guidance of neuronal growth cones and apoptosis,^94^ and has been associated with suicide traits^18^ and multiple psychiatric disorders, as reported in the GWAS Catalog.^59^ TWAS highlighted cerebellar tissue, which showed a high number of significant genes, corroborating the enrichment patterns observed in MAGMA gene expression analysis. Frontal cortex BA9 and basal ganglia (putamen and caudate) tissues were also highlighted, suggesting potential contributions of cortico-striatal circuits to suicide vulnerability.^100,101^ In addition to brain-derived tissues, transcriptional regulation in non-brain body tissues (e.g., peripheral blood^102^) may also contribute to suicide risk. Our isoTWAS analysis in particular identified additional genes (e.g., *CLYBL*, *THAP3*, *SPG7*, *ZNF530*, *SIPA1L3*) compared to TWAS, despite using only frontal cortex tissue. CpG sites and mapped genes identified from mQTL-SMR implicated that DNA methylation variation may contribute to suicide neuropathology.^103^ PWAS identified only one protein (CTNND1) shared between SI and SA, further supporting that the molecular basis of these suicide traits only partially overlap.^20^ Integrating prioritization results from all five approaches across the three suicide traits, *UGGT2*, *GMPPB*, and *BRWD1* were genes with the highest score of 8. *UGGT2* showed convergent evidence for all three suicide traits. It encodes a UDP-glucose:glycoprotein glucosyltransferase that provides quality control for protein folding in the endoplasmic reticulum^94^ and has been associated with schizophrenia and Alzheimer’s disease.^59^ *GMPPB* and *BRWD1* showed evidence for both SA and SP. *GMPPB* encodes a GDP-mannose pyrophosphorylase involved in the production of N-linked oligosaccharides^94^ and has been associated with Alzheimer’s disease.^59^ *BRWD1* encodes a protein involved in the regulation of cell morphology and cytoskeletal organization^94^ and has been associated with substance use disorders and ADHD, autism spectrum disorder, and Alzheimer’s disease.^59^ Among the 16 genes shared across the three suicide traits, most are involved in neuronal and synaptic functions (*DCC*, *RBFOX1*, *GRIA1*, *ANKK1*, *CTNND1*, *KTN1-AS1*, *STYXL1*). Additional groups included genes related to metabolic and mitochondrial pathways (*MDH2*, *SFXN2*, *CLYBL*, *ARL3*, *METAP1D*), protein processing (*UGGT2*, *ALG12*, *DNAJC24*), and immune regulation (*PAX5*).^94^ Comparing genes exclusively associated with SI and SA, SA-associated genes tended to be implicated in synaptic structure and neuronal signaling, whereas SI-associated genes appeared to be more strongly related to transcriptional regulation, intracellular processing, and cellular stress-response, with more modest neuronal involvement. Although most gene-discovery signals were related to non-coding regulatory variants, we also observed supporting evidence from coding variants. For example, missense variants in *ANKK1* and *SYCE1L*, and synonymous variants in *MON1B*, *PSMG1*, and *DRD2*, all showed significant association signals in suicide GWASs, and these genes also had high prioritization scores. This implies the effects of protein-structure alterations and potential functional consequences of “silent” synonymous coding changes on suicide risk.^104^

GSA-MiXeR gene-set analysis revealed differences in GO term enrichment between suicide traits. For example, SA, but not SI, showed heritability enrichment in apoptotic process involved in development, suggesting that programmed cell death may be relevant to SA and its associated risks.^105^ We also observed convergent GO term enrichment across the three suicide traits, such as cellular response to stress, Golgi apparatus, and transcription regulator activity, which have been previously linked to psychological stress,^106^ neurodegenerative^107^ and psychiatric disorders.^108^ In drug repurposing analysis using DRUGSETS, we identified five drug candidates for SA and SP, all of which are antipsychotics, highlighting the importance of managing psychiatric disorders in suicide prevention^109^ and pointing to potential therapeutic targets involving topoisomerase inhibitor and sodium channel blocker pathways.

Both the significant genetic correlations between suicide traits and psychiatric disorders and the persistence of SNP-based heritability after conditioning on psychiatric disorders support the mediating role of psychiatric disorders in the liability to suicidality.^110^ However, the effects varied across these disorders. For SA, ANX, MDD, and PTSD all showed a high genetic correlation, and conditioning on ANX produced the largest reduction in heritability; but for SI, both the highest genetic correlation and heritability reduction were observed for MDD. This suggested that SA and SI may be predominantly driven by different psychopathologies.^16,20^ Nevertheless, although attenuated after conditioning, the residual heritability of suicide traits and hundreds of genetic correlations retained significance, indicating the presence of genetic mechanisms of suicide traits partially independent of these psychiatric conditions.^15^ In MiXeR analysis, some trait pairs (SCZ–SI, SUD–SI, and BIP–SA) showed relatively low genetic correlations but a high proportion of shared SNPs, implying substantial overlap in the underlying polygenic architecture between these traits, despite heterogeneity in effect directions among shared variants.^49^

We observed consistent evidence supporting potential causal effects of three factors on suicidal behaviors. Social isolation and loneliness were significant predictors of both SI and SA, with depression potentially acting as a mediator, whereas social support exhibited a protective effect on suicide risk.^111^ We also identified a causal effect of medical abortion. However, this association remains controversial in the literature, with some studies reporting associations between abortion histories and SA^112^ while others attributing this to confounding factors rather than a direct causal link.^113^ Cross-sectional studies similarly reported elevated suicidal behaviors in those with early sexual initiation,^114^ and our analyses provided complementary genetic evidence further supporting this relationship. In addition to risk factors, convergent evidence also indicated that suicidal behaviors may lead to a range of health-related consequences. These outcomes were predominantly physical conditions, including spinal stenosis, knee internal derangement, essential hypertension, and difficulty falling asleep, as well as multiple drug-use-related mental disorders and hospital admissions. SA may directly result in injuries requiring medical treatment and hospitalization,^115^ and may also contribute to alterations in brain structure and exacerbate existing psychiatric conditions, thereby increasing vulnerability to diverse adverse outcomes later in life.^116,117^ The observed association between suicide traits and parental phenotypes may reflect dynastic effects,^118^ wherein intergenerationally shared genetic risks influence both parental and offspring characteristics through shared family environments.

In addition to the putative causal relationships described above, our genetic correlation analysis revealed a broader spectrum of correlated domains, ranging from biomarkers to clinical and behavioral traits, and extending to more distal socioeconomic factors. Beyond mental health and physical health-related traits, we also identified correlations within the behavioral domain, involving substance use, sleep patterns, physical activity, and dietary habits.^5,21–23^ As modifiable factors, these risk behaviors may be emphasized in health promotion and suicide prevention efforts. We additionally observed correlations with brain structure and function measures,^98^ including reduced volumes in multiple cerebellar regions, consistent with our gene-discovery findings. Metabolomic analysis further highlighted associations related to phospholipid levels as well as monounsaturated and polyunsaturated fatty acids.^119^ Moreover, we identified distal correlated factors, such as material deprivation, low socioeconomic status, and limited social relationships, which may interact with mental health, physical health, and behavioral traits. Their causal pathways need to be disentangled in future studies.^120^

The present study has several strengths. First, by integrating ancestrally diverse cohorts, we identified 90 independent lead SNPs for suicide traits, 49 of which were not previously reported. We identified lead SNPs in non-EUR ancestries, representing the first report of genomic associations for SI in some population groups. Second, we applied multiple complementary gene-discovery approaches and prioritized 1052 genes, most of which were reported for the first time. Third, in addition to those with psychiatric disorders, we identified putative causal associations between phenome-wide complex traits and suicide, as supported by convergent evidence from multiple approaches. However, this study also has limitations. Although EHR-based suicide assessment is generally more precise than the self-reported, suicide history among at least a subset of EHR-based controls may be unknown, which is likely to underestimate suicide prevalence, introduce case–control misclassification, and attenuate GWAS effect sizes. In addition, there was heterogeneity in suicide definitions across cohorts, as different survey questions or diagnostic criteria in EHR data were applied. For example, in FinnGen, SA cases defined using ICD-10 codes X60-X84 can include non-suicidal self-harm. Some cohorts in PGC GWAS were designed to investigate specific psychiatric disorders,^17^ and their associations may therefore differ from those observed in general populations, although we partially mitigated this by including GWAS performed in community-based cohorts such as UKB, AoU, and FinnGen. There was also potential sample overlap between cohorts, for example between AoU and MVP, which should ideally be identified and excluded through collaboration between cohorts. Finally, for non-EUR ancestries, limited sample sizes resulted in insufficient power to estimate SNP-based heritability and genetic correlations, which precluded the use of gSEM to characterize their multivariate genetic architecture.

In conclusion, the present study provides genetic evidence for suicidal behaviors at both SNP and gene levels and reveals relevant risk factors and consequences at a phenome-wide scale. These findings deepen our understanding of the molecular mechanisms underlying the development of suicidality and inform strategies for suicide control and prevention in both clinical and public health settings. Future work can build on this investigation to further characterize the genetic architecture of suicidal behaviors focusing on rare and structural variants, establish higher-resolution gene-prioritization map applying single-cell and spatial transcriptomic approaches, and delineate causal networks of suicidal behaviors that integrate biological, psychosocial, and socioeconomic factors.

## Data Availability

The GWAS summary statistics generated in the present study will be made publicly available online upon publication. Individual-level data were obtained from UKB (http://www.ukbiobank.ac.uk/) and the AoU Curated Data Repository version 8 (https://www.researchallofus.org). GWAS data used in this study were downloaded from Pan-UKB (https://pan.ukbb.broadinstitute.org/), FinnGen (https://www.finngen.fi/en/access_results), MVP (dbGaP accession numbers phs001672 and phs002453; https://dbgap.ncbi.nlm.nih.gov/), and PGC (https://www.med.unc.edu/pgc/download-results/).

## Contributors

JH and RP conceived and designed the study. JH, BCM, DQ, DD, ZM, QC, EP, QZ, and SK conducted the formal analysis and contributed to the data interpretation. JH drafted the manuscript. All authors participated in the critical revision and editing of the manuscript. JH obtained the primary funding, and RP supervised the study. All authors read and approved the final version of the manuscript.

## Acknowledgments

This study was funded by the American Foundation for Suicide Prevention (PDF-0-065-23 to JH). RP acknowledges support from the National Institute of Mental Health (RF1MH132337). BCM acknowledges support from MQ: Transforming Mental Health (UFA21\100014). The authors sincerely thank all the participants enrolled in UKB, AoU, FinnGen, MVP, and PGC, as well as the investigators contributing to these cohorts. We also gratefully acknowledge UKB and the National Institutes of Health’s AoU for making available the individual-level participant data examined in this study. The research using UKB resources has been conducted under Application Number 58146. The AoU Research Program is supported by the National Institutes of Health, Office of the Director: Regional Medical Centers: 1 OT2 OD026549; 1 OT2 OD026554; 1 OT2 OD026557; 1 OT2 OD026556; 1OT2 OD026550; 1OT2OD026552; 1OT2 OD026553; 1 OT2 OD026548; 1 OT2 OD026551; 1 OT2 OD026555; IAA #: AOD 16037; Federally Qualified Health Centers: HHSN 263201600085U; Data and Research Center: 5 U2C OD023196; Biobank: 1 U24 OD023121; The Participant Center: U24 OD023176; Participant Technology Systems Center: 1 U24 OD023163; Communications and Engagement: 3 OT2 OD023205; 3 OT2 OD023206; and Community Partners: 1 OT2 OD025277; 3 OT2 OD025315; 1 OT2 OD025337; 1 OT2OD025276.

## References

1. GBD 2023 Causes of Death Collaborators. Global burden of 292 causes of death in 204 countries and territories and 660 subnational locations, 1990-2023: a systematic analysis for the Global Burden of Disease Study 2023. Lancet 2025; 406(10513): 1811–72.

2. Cabrera-Mendoza B, Fries GR, Polimanti R, Suicide Working Group of the Latin American Genomics C. The landscape of suicide risk factors in Latin America. Psychiatry Res 2025; 351: 116597.

3. GBD 2021 Suicide Collaborators. Global, regional, and national burden of suicide, 1990-2021: a systematic analysis for the Global Burden of Disease Study 2021. Lancet Public Health 2025; 10(3): e189–e202.

4. He J, Ouyang F, Qiu D, Li L, Li Y, Xiao S. Time Trends and Predictions of Suicide Mortality for People Aged 70 Years and Over From 1990 to 2030 Based on the Global Burden of Disease Study 2017. Front Psychiatry 2021; 12: 721343.

5. Favril L, Yu R, Geddes JR, Fazel S. Individual-level risk factors for suicide mortality in the general population: an umbrella review. Lancet Public Health 2023; 8(11): e868–e77.

6. Goldman-Mellor SJ, Caspi A, Harrington H, et al. Suicide attempt in young people: a signal for long-term health care and social needs. JAMA Psychiatry 2014; 71(2): 119–27.

7. Peterson C, Haileyesus T, Stone DM. Economic Cost of U.S. Suicide and Nonfatal Self-harm. Am J Prev Med 2024; 67(1): 129–33.

8. Nigam A, Vuddemarry M, Zadey S. Economic burden of suicide deaths in India (2019): a retrospective, cross-sectional study. Lancet Reg Health Southeast Asia 2024; 29: 100477.

9. Mullins N, Bigdeli TB, Borglum AD, et al. GWAS of Suicide Attempt in Psychiatric Disorders and Association With Major Depression Polygenic Risk Scores. Am J Psychiatry 2019; 176(8): 651–60.

10. Strawbridge RJ, Ward J, Ferguson A, et al. Identification of novel genome-wide associations for suicidality in UK Biobank, genetic correlation with psychiatric disorders and polygenic association with completed suicide. EBioMedicine 2019; 41: 517–25.

11. Docherty AR, Shabalin AA, DiBlasi E, et al. Genome-Wide Association Study of Suicide Death and Polygenic Prediction of Clinical Antecedents. Am J Psychiatry 2020; 177(10): 917–27.

12. Erlangsen A, Appadurai V, Wang Y, et al. Genetics of suicide attempts in individuals with and without mental disorders: a population-based genome-wide association study. Mol Psychiatry 2020; 25(10): 2410–21.

13. Ruderfer DM, Walsh CG, Aguirre MW, et al. Significant shared heritability underlies suicide attempt and clinically predicted probability of attempting suicide. Mol Psychiatry 2020; 25(10): 2422–30.

14. Kimbrel NA, Ashley-Koch AE, Qin XJ, et al. A genome-wide association study of suicide attempts in the million veterans program identifies evidence of pan-ancestry and ancestry-specific risk loci. Mol Psychiatry 2022; 27(4): 2264–72.

15. Mullins N, Kang J, Campos AI, et al. Dissecting the Shared Genetic Architecture of Suicide Attempt, Psychiatric Disorders, and Known Risk Factors. Biol Psychiatry 2022; 91(3): 313–27.

16. Ashley-Koch AE, Kimbrel NA, Qin XJ, et al. Genome-wide association study identifies four pan-ancestry loci for suicidal ideation in the Million Veteran Program. PLoS Genet 2023; 19(3): e1010623.

17. Docherty AR, Mullins N, Ashley-Koch AE, et al. GWAS Meta-Analysis of Suicide Attempt: Identification of 12 Genome-Wide Significant Loci and Implication of Genetic Risks for Specific Health Factors. Am J Psychiatry 2023; 180(10): 723–38.

18. Kimbrel NA, Ashley-Koch AE, Qin XJ, et al. Identification of Novel, Replicable Genetic Risk Loci for Suicidal Thoughts and Behaviors Among US Military Veterans. JAMA Psychiatry 2023; 80(2): 135–45.

19. Li QS, Shabalin AA, DiBlasi E, et al. Genome-wide association study meta-analysis of suicide death and suicidal behavior. Mol Psychiatry 2023; 28(2): 891–900.

20. Colbert SMC, Group tPGCSW, Ruderfer D, Docherty AR, Mullins N. Genome-wide association studies identify 77 loci for suicidality and provide novel biological insights. medRxiv 2025: 2025.10.22.25338076.

21. Turecki G, Brent DA, Gunnell D, et al. Suicide and suicide risk. Nat Rev Dis Primers 2019; 5(1): 74.

22. Fazel S, Runeson B. Suicide. N Engl J Med 2020; 382(3): 266–74.

23. Favril L, Yu R, Uyar A, Sharpe M, Fazel S. Risk factors for suicide in adults: systematic review and meta-analysis of psychological autopsy studies. Evid Based Ment Health 2022; 25(4): 148–55.

24. Batty GD, Kivimaki M, Bell S, et al. Psychosocial characteristics as potential predictors of suicide in adults: an overview of the evidence with new results from prospective cohort studies. Transl Psychiatry 2018; 8(1): 22.

25. Franklin JC, Ribeiro JD, Fox KR, et al. Risk factors for suicidal thoughts and behaviors: A meta-analysis of 50 years of research. Psychol Bull 2017; 143(2): 187–232.

26. Pingault JB, Richmond R, Davey Smith G. Causal Inference with Genetic Data: Past, Present, and Future. Cold Spring Harb Perspect Med 2022; 12(3).

27. Bycroft C, Freeman C, Petkova D, et al. The UK Biobank resource with deep phenotyping and genomic data. Nature 2018; 562(7726): 203–9.

28. All of Us Research Program Investigators, Denny JC, Rutter JL, et al. The “All of Us” Research Program. N Engl J Med 2019; 381(7): 668–76.

29. Gaziano JM, Concato J, Brophy M, et al. Million Veteran Program: A mega-biobank to study genetic influences on health and disease. J Clin Epidemiol 2016; 70: 214–23.

30. Kurki MI, Karjalainen J, Palta P, et al. FinnGen provides genetic insights from a well-phenotyped isolated population. Nature 2023; 613(7944): 508–18.

31. Agrawal A, Bulik CM, Abebe DS, et al. The Psychiatric Genomics Consortium: discoveries and directions. Lancet Psychiatry 2025; 12(8): 600–10.

32. Sullivan PF, Agrawal A, Bulik CM, et al. Psychiatric Genomics: An Update and an Agenda. Am J Psychiatry 2018; 175(1): 15–27.

33. Grotzinger AD, Rhemtulla M, de Vlaming R, et al. Genomic structural equation modelling provides insights into the multivariate genetic architecture of complex traits. Nat Hum Behav 2019; 3(5): 513–25.

34. Willer CJ, Li Y, Abecasis GR. METAL: fast and efficient meta-analysis of genomewide association scans. Bioinformatics 2010; 26(17): 2190–1.

35. Watanabe K, Taskesen E, van Bochoven A, Posthuma D. Functional mapping and annotation of genetic associations with FUMA. Nat Commun 2017; 8(1): 1826.

36. Yang J, Ferreira T, Morris AP, et al. Conditional and joint multiple-SNP analysis of GWAS summary statistics identifies additional variants influencing complex traits. Nat Genet 2012; 44(4): 369–75, S1-3.

37. Yuan K, Longchamps RJ, Pardinas AF, et al. Fine-mapping across diverse ancestries drives the discovery of putative causal variants underlying human complex traits and diseases. Nat Genet 2024; 56(9): 1841–50.

38. de Leeuw CA, Mooij JM, Heskes T, Posthuma D. MAGMA: generalized gene-set analysis of GWAS data. PLoS Comput Biol 2015; 11(4): e1004219.

39. Barbeira AN, Dickinson SP, Bonazzola R, et al. Exploring the phenotypic consequences of tissue specific gene expression variation inferred from GWAS summary statistics. Nat Commun 2018; 9(1): 1825.

40. Barbeira AN, Pividori M, Zheng J, Wheeler HE, Nicolae DL, Im HK. Integrating predicted transcriptome from multiple tissues improves association detection. PLoS Genet 2019; 15(1): e1007889.

41. Bhattacharya A, Vo DD, Jops C, et al. Isoform-level transcriptome-wide association uncovers genetic risk mechanisms for neuropsychiatric disorders in the human brain. Nat Genet 2023; 55(12): 2117–28.

42. Gusev A, Ko A, Shi H, et al. Integrative approaches for large-scale transcriptome-wide association studies. Nat Genet 2016; 48(3): 245–52.

43. Wu Y, Zeng J, Zhang F, et al. Integrative analysis of omics summary data reveals putative mechanisms underlying complex traits. Nat Commun 2018; 9(1): 918.

44. Zhu Z, Zhang F, Hu H, et al. Integration of summary data from GWAS and eQTL studies predicts complex trait gene targets. Nat Genet 2016; 48(5): 481–7.

45. Frei O, Hindley G, Shadrin AA, et al. Improved functional mapping of complex trait heritability with GSA-MiXeR implicates biologically specific gene sets. Nat Genet 2024; 56(6): 1310–8.

46. Bell N, Uffelmann E, van Walree E, de Leeuw C, Posthuma D. Using genome-wide association results to identify drug repurposing candidates. medRxiv 2022.

47. Bulik-Sullivan BK, Loh PR, Finucane HK, et al. LD Score regression distinguishes confounding from polygenicity in genome-wide association studies. Nat Genet 2015; 47(3): 291–5.

48. Bulik-Sullivan B, Finucane HK, Anttila V, et al. An atlas of genetic correlations across human diseases and traits. Nat Genet 2015; 47(11): 1236–41.

49. Frei O, Holland D, Smeland OB, et al. Bivariate causal mixture model quantifies polygenic overlap between complex traits beyond genetic correlation. Nat Commun 2019; 10(1): 2417.

50. O’Connor LJ, Price AL. Distinguishing genetic correlation from causation across 52 diseases and complex traits. Nat Genet 2018; 50(12): 1728–34.

51. Mounier N, Kutalik Z. Bias correction for inverse variance weighting Mendelian randomization. Genet Epidemiol 2023; 47(4): 314–31.

52. Xue A, Zhu Z, Wang H, et al. Unravelling the complex causal effects of substance use behaviours on common diseases. Commun Med (Lond) 2024; 4(1): 43.

53. All of Us Research Program Genomics Investigators. Genomic data in the All of Us Research Program. Nature 2024; 627(8003): 340–6.

54. Karczewski KJ, Gupta R, Kanai M, et al. Pan-UK Biobank genome-wide association analyses enhance discovery and resolution of ancestry-enriched effects. Nat Genet 2025; 57(10): 2408–17.

55. Chang CC, Chow CC, Tellier LC, Vattikuti S, Purcell SM, Lee JJ. Second-generation PLINK: rising to the challenge of larger and richer datasets. Gigascience 2015; 4: 7.

56. Furtjes AE, Arathimos R, Coleman JRI, et al. General dimensions of human brain morphometry inferred from genome-wide association data. Hum Brain Mapp 2023; 44(8): 3311–23.

57. International HapMap Consortium, Altshuler DM, Gibbs RA, et al. Integrating common and rare genetic variation in diverse human populations. Nature 2010; 467(7311): 52–8.

58. Genomes Project Consortium, Auton A, Brooks LD, et al. A global reference for human genetic variation. Nature 2015; 526(7571): 68–74.

59. Cerezo M, Sollis E, Ji Y, et al. The NHGRI-EBI GWAS Catalog: standards for reusability, sustainability and diversity. Nucleic Acids Res 2025; 53(D1): D998–D1005.

60. Yang J, Lee SH, Goddard ME, Visscher PM. GCTA: a tool for genome-wide complex trait analysis. Am J Hum Genet 2011; 88(1): 76–82.

61. McLaren W, Gil L, Hunt SE, et al. The Ensembl Variant Effect Predictor. Genome Biol 2016; 17(1): 122.

62. GTEx Consortium. The GTEx Consortium atlas of genetic regulatory effects across human tissues. Science 2020; 369(6509): 1318–30.

63. PredictDB Team. PredictDB Data Repository. https://predictdb.org/ (accessed July 1 2025).

64. Bhattacharya A. isoTWAS models using 48 GTEx tissues and PsychENCODE data. 2022. https://zenodo.org/record/6795947#.Y8mi2-zMLBI (accessed June 17 2025).

65. Beach TG, Adler CH, Sue LI, et al. Arizona Study of Aging and Neurodegenerative Disorders and Brain and Body Donation Program. Neuropathology 2015; 35(4): 354–89.

66. Beach TG, Sue LI, Walker DG, et al. The Sun Health Research Institute Brain Donation Program: description and experience, 1987-2007. Cell Tissue Bank 2008; 9(3): 229–45.

67. Qi T, Wu Y, Zeng J, et al. Identifying gene targets for brain-related traits using transcriptomic and methylomic data from blood. Nat Commun 2018; 9(1): 2282.

68. Ng B, White CC, Klein HU, et al. An xQTL map integrates the genetic architecture of the human brain’s transcriptome and epigenome. Nat Neurosci 2017; 20(10): 1418–26.

69. Hannon E, Spiers H, Viana J, et al. Methylation QTLs in the developing brain and their enrichment in schizophrenia risk loci. Nat Neurosci 2016; 19(1): 48–54.

70. Jaffe AE, Gao Y, Deep-Soboslay A, et al. Mapping DNA methylation across development, genotype and schizophrenia in the human frontal cortex. Nat Neurosci 2016; 19(1): 40–7.

71. Min JL, Hemani G, Hannon E, et al. Genomic and phenotypic insights from an atlas of genetic effects on DNA methylation. Nat Genet 2021; 53(9): 1311–21.

72. Gaunt TR, Shihab HA, Hemani G, et al. Systematic identification of genetic influences on methylation across the human life course. Genome Biol 2016; 17: 61.

73. Hatton AA, Cheng FF, Lin T, et al. Genetic control of DNA methylation is largely shared across European and East Asian populations. Nat Commun 2024; 15(1): 2713.

74. McRae AF, Marioni RE, Shah S, et al. Identification of 55,000 Replicated DNA Methylation QTL. Sci Rep 2018; 8(1): 17605.

75. Hannon E, Dempster E, Viana J, et al. An integrated genetic-epigenetic analysis of schizophrenia: evidence for co-localization of genetic associations and differential DNA methylation. Genome Biol 2016; 17(1): 176.

76. Hannon E, Gorrie-Stone TJ, Smart MC, et al. Leveraging DNA-Methylation Quantitative-Trait Loci to Characterize the Relationship between Methylomic Variation, Gene Expression, and Complex Traits. Am J Hum Genet 2018; 103(5): 654–65.

77. Sayols S. rrvgo: a Bioconductor package for interpreting lists of Gene Ontology terms. MicroPubl Biol 2023; 2023.

78. Major Depressive Disorder Working Group of the Psychiatric Genomics Consortium. Trans-ancestry genome-wide study of depression identifies 697 associations implicating cell types and pharmacotherapies. Cell 2025; 188(3): 640–52 e9.

79. Nievergelt CM, Maihofer AX, Atkinson EG, et al. Genome-wide association analyses identify 95 risk loci and provide insights into the neurobiology of post-traumatic stress disorder. Nat Genet 2024; 56(5): 792–808.

80. Friligkou E, Lokhammer S, Cabrera-Mendoza B, et al. Gene discovery and biological insights into anxiety disorders from a large-scale multi-ancestry genome-wide association study. Nat Genet 2024; 56(10): 2036–45.

81. O’Connell KS, Koromina M, van der Veen T, et al. Genomics yields biological and phenotypic insights into bipolar disorder. Nature 2025; 639(8056): 968–75.

82. Trubetskoy V, Pardinas AF, Qi T, et al. Mapping genomic loci implicates genes and synaptic biology in schizophrenia. Nature 2022; 604(7906): 502–8.

83. Hatoum AS, Colbert SMC, Johnson EC, et al. Multivariate genome-wide association meta-analysis of over 1 million subjects identifies loci underlying multiple substance use disorders. Nat Ment Health 2023; 1(3): 210–23.

84. Zhu Z, Zheng Z, Zhang F, et al. Causal associations between risk factors and common diseases inferred from GWAS summary data. Nat Commun 2018; 9(1): 224.

85. Verma A, Huffman JE, Rodriguez A, et al. Diversity and scale: Genetic architecture of 2068 traits in the VA Million Veteran Program. Science 2024; 385(6706).

86. IEU Open GWAS Project. Metabolic biomarkers in the UK Biobank measured by Nightingale Health 2020. 2020. https://gwas.mrcieu.ac.uk/datasets/?gwas_id_icontains=met-d (accessed June 1 2025).

87. Karjalainen MK, Karthikeyan S, Oliver-Williams C, et al. Genome-wide characterization of circulating metabolic biomarkers. Nature 2024; 628(8006): 130–8.

88. Smith SM, Douaud G, Chen W, et al. An expanded set of genome-wide association studies of brain imaging phenotypes in UK Biobank. Nat Neurosci 2021; 24(5): 737–45.

89. Cross-Disorder Group of the Psychiatric Genomics Consortium. Genomic Relationships, Novel Loci, and Pleiotropic Mechanisms across Eight Psychiatric Disorders. Cell 2019; 179(7): 1469–82 e11.

90. Peyrot WJ, Price AL. Identifying loci with different allele frequencies among cases of eight psychiatric disorders using CC-GWAS. Nat Genet 2021; 53(4): 445–54.

91. Shin JJH, Gillingham AK, Begum F, Chadwick J, Munro S. TBC1D23 is a bridging factor for endosomal vesicle capture by golgins at the trans-Golgi. Nat Cell Biol 2017; 19(12): 1424–32.

92. Sinnott-Armstrong N, Tanigawa Y, Amar D, et al. Genetics of 35 blood and urine biomarkers in the UK Biobank. Nat Genet 2021; 53(2): 185–94.

93. Simard M, Madore AM, Girard S, et al. Polygenic risk score for atopic dermatitis in the Canadian population. J Allergy Clin Immunol 2021; 147(1): 406–9.

94. GeneCards®: The Human Gene Database. https://www.genecards.org/.

95. Wendt FR, Pathak GA, Deak JD, et al. Using phenotype risk scores to enhance gene discovery for generalized anxiety disorder and posttraumatic stress disorder. Mol Psychiatry 2022; 27(4): 2206–15.

96. Chen Y, Liu P, Yi S, Fan C, Zhao W, Liu J. Investigating the shared genetic architecture between attention-deficit/hyperactivity disorder and risk taking behavior: A large-scale genomewide cross-trait analysis. J Affect Disord 2024; 356: 22–31.

97. Ikeda M, Takahashi A, Kamatani Y, et al. Genome-Wide Association Study Detected Novel Susceptibility Genes for Schizophrenia and Shared Trans-Populations/Diseases Genetic Effect. Schizophr Bull 2019; 45(4): 824–34.

98. Dobbertin M, Blair KS, Carollo E, Blair JR, Dominguez A, Bajaj S. Neuroimaging alterations of the suicidal brain and its relevance to practice: an updated review of MRI studies. Front Psychiatry 2023; 14: 1083244.

99. Keaton SA, Madaj ZB, Heilman P, et al. An inflammatory profile linked to increased suicide risk. J Affect Disord 2019; 247: 57–65.

100. Pu S, Nakagome K, Yamada T, et al. Suicidal ideation is associated with reduced prefrontal activation during a verbal fluency task in patients with major depressive disorder. J Affect Disord 2015; 181: 9–17.

101. Bowden C, Cheetham SC, Lowther S, Katona CL, Crompton MR, Horton RW. Reduced dopamine turnover in the basal ganglia of depressed suicides. Brain Res 1997; 769(1): 135–40.

102. Sun S, Liu Q, Wang Z, et al. Brain and blood transcriptome profiles delineate common genetic pathways across suicidal ideation and suicide. Mol Psychiatry 2024; 29(5): 1417–26.

103. Policicchio S, Washer S, Viana J, et al. Genome-wide DNA methylation meta-analysis in the brains of suicide completers. Transl Psychiatry 2020; 10(1): 69.

104. Kaissarian NM, Meyer D, Kimchi-Sarfaty C. Synonymous Variants: Necessary Nuance in Our Understanding of Cancer Drivers and Treatment Outcomes. J Natl Cancer Inst 2022; 114(8): 1072–94.

105. McKernan DP, Dinan TG, Cryan JF. “Killing the Blues”: a role for cellular suicide (apoptosis) in depression and the antidepressant response? Prog Neurobiol 2009; 88(4): 246–63.

106. Hayashi T. Conversion of psychological stress into cellular stress response: roles of the sigma-1 receptor in the process. Psychiatry Clin Neurosci 2015; 69(4): 179–91.

107. Rabouille C, Haase G. Editorial: Golgi Pathology in Neurodegenerative Diseases. Front Neurosci 2015; 9: 489.

108. Cabrera-Mendoza B, de Anda-Jauregui G, Nicolini H, Fresno C. A meta-study on transcription factor networks in the suicidal brain. J Psychiatr Res 2021; 136: 23–31.

109. Hassan A, De Luca V, Dai N, et al. Effectiveness of Antipsychotics in Reducing Suicidal Ideation: Possible Physiologic Mechanisms. Healthcare (Basel) 2021; 9(4).

110. Hoertel N, Franco S, Wall MM, et al. Mental disorders and risk of suicide attempt: a national prospective study. Mol Psychiatry 2015; 20(6): 718–26.

111. Motillon-Toudic C, Walter M, Seguin M, Carrier JD, Berrouiguet S, Lemey C. Social isolation and suicide risk: Literature review and perspectives. Eur Psychiatry 2022; 65(1): e65.

112. Reardon DC. Suicide risks associated with pregnancy outcomes: a national cross-sectional survey of American females 41-45 years of age. J Psychosom Obstet Gynaecol 2025; 46(1): 2455086.

113. Steinberg JR, Laursen TM, Adler NE, Gasse C, Agerbo E, Munk-Olsen T. The association between first abortion and first-time non-fatal suicide attempt: a longitudinal cohort study of Danish population registries. Lancet Psychiatry 2019; 6(12): 1031–8.

114. Ren J, Qi X, Cao W, et al. Early Sexual Initiation Is Associated with Suicide Attempts among Chinese Young People. Int J Environ Res Public Health 2022; 19(7).

115. Rocos B, Acharya M, Chesser TJ. The Pattern of Injury and Workload Associated with Managing Patients After Suicide Attempt by Jumping from a Height. Open Orthop J 2015; 9: 395–8.

116. Auger N, Chadi N, Ayoub A, Brousseau E, Low N. Suicide Attempt and Risk of Substance Use Disorders Among Female Youths. JAMA Psychiatry 2022; 79(7): 710–7.

117. Shanahan L, Schorpp KM, Volpe VV, Linthicum K, Freeman JA. Developmental timing of suicide attempts and cardiovascular risk during young adulthood. Health Psychol 2016; 35(10): 1135–43.

118. Hwang LD, Davies NM, Warrington NM, Evans DM. Integrating Family-Based and Mendelian Randomization Designs. Cold Spring Harb Perspect Med 2021; 11(3).

119. Sublette ME, Hibbeln JR, Galfalvy H, Oquendo MA, Mann JJ. Omega-3 polyunsaturated essential fatty acid status as a predictor of future suicide risk. Am J Psychiatry 2006; 163(6): 1100–2.

120. Na PJ, Shin J, Kwak HR, et al. Social Determinants of Health and Suicide-Related Outcomes: A Review of Meta-Analyses. JAMA Psychiatry 2025; 82(4): 337–46.

