## appendix for "Multivariate genome-wide association study of suicidal behaviors in >1.7 million individuals of diverse population descents"

**Supplemental tables**

Detailed information can be found in Supplemental tables.xlsx.

**Contents:**


**Table S1A.** Sources of GWAS for suicide phenotypes across individual cohorts.
**Table S1B.** Source data for suicide ideation and suicide attempt definitions in UK Biobank (UKB) and All of Us Research Program (AoU).
**Table S2A.** SNP-based heritability of suicide phenotypes in European descent before and after mtCOJO conditioning on six psychiatric disorders.
**Table S2B.** Genetic correlation of suicide phenotypes across cohorts in European ancestry.
**Table S3A.** Lead SNPs and significant genomic loci associated with suicidality spectrum (SP) in European descent and those exclusive for suicide ideation (SI) and suicide attempt (SA).
**Table S3B.** GWAS Catalog traits previously associated with lead SNPs identified in this study.
**Table S3C.** Lead SNPs and significant genomic loci associated with suicide ideation in each ancestry.
**Table S3D.** Lead SNPs and significant genomic loci associated with suicide attempt in each ancestry.
**Table S3E.** Cross-ancestry meta-analysis and ancestry-exclusive lead SNPs and significant genomic loci associated with suicide ideation.
**Table S3F.** Cross-ancestry meta-analysis and ancestry-exclusive lead SNPs and significant genomic loci associated with suicide attempt.
**Table S4.** Additional independent genome-wide significant SNPs identified by COJO analysis in EUR descent.
**Table S5.** Credible sets and putative causal SNPs for suicide phenotypes identified by SuSiEx fine-mapping of multi-ancestry GWAS.
**Table S6.** Significant eQTL-based tissue enrichments from MAGMA expression analysis of suicide phenotypes across ancestries.
**Table S7.** Significant MAGMA gene-based associations for suicide phenotypes across ancestries.
**Table S8A.** Genetically regulated transcriptomic associations with suicide ideation identified by cross-tissue TWAS using MetaXcan.
**Table S8B.** Genetically regulated transcriptomic associations with suicide attempt identified by cross-tissue TWAS using MetaXcan.
**Table S8C.** Genetically regulated transcriptomic associations with suicidality spectrum identified by cross-tissue TWAS using MetaXcan.
**Table S9A.** FDR-significant isoform-level transcriptome-wide associations with suicide ideation in adult frontal cortex.
**Table S9B.** FDR-significant isoform-level transcriptome-wide associations with suicide attempt in adult frontal cortex.
**Table S9C.** FDR-significant isoform-level transcriptome-wide associations with suicidality spectrum in adult frontal cortex.

**Table S10.** Genetically predicted brain proteins in dorsolateral prefrontal cortex for suicide phenotypes in European descent identified by PWAS using FUSION.
**Table S11A.** CpG islands associated with suicide ideation identified by SMR analysis.
**Table S11B.** CpG islands associated with suicide attempt identified by SMR analysis.
**Table S11C.** CpG islands associated with suicidality spectrum identified by SMR analysis.
**Table S12.** Summary of genes associated with suicide phenotypes across five complementary approaches.
**Table S13A.** Gene sets associated with suicide phenotypes based on Gene Ontology (GO) fold-enrichment estimates from GSA-MiXeR.
**Table S13B.** Reduced Gene Ontology (GO) terms associated with suicide phenotypes identified using rrvgo.
**Table S14.** Drug repurposing candidates for suicide phenotypes from drug gene set analysis (DRUGSETS).
**Table S15.** Genetic correlation between suicide phenotypes and psychiatric disorders.
**Table S16.** Causal variants shared between suicide phenotypes and psychiatric disorders estimated by bivariate MiXeR model.
**Table S17.** Phenome-wide genetic correlations (rg) between suicide phenotypes and traits from UK Biobank (UKB), FinnGen, Million Veteran Program (MVP), metabolomics, and brain image-derived phenotypes (IDP), before and after mtCOJO conditioning on six psychiatric disorders.
**Table S18.** Significant putative causal effects between suicide phenotypes and phenome-wide traits using latent causal variable analysis.
**Table S19.** Putative causal associations between suicide phenotypes and phenome-wide traits estimated by bidirectional Mendelian randomization using MRlap.
**Table S20.** Putative causal associations between suicide phenotypes and phenome-wide traits identified using GSMR2 in non-overlapping datasets.

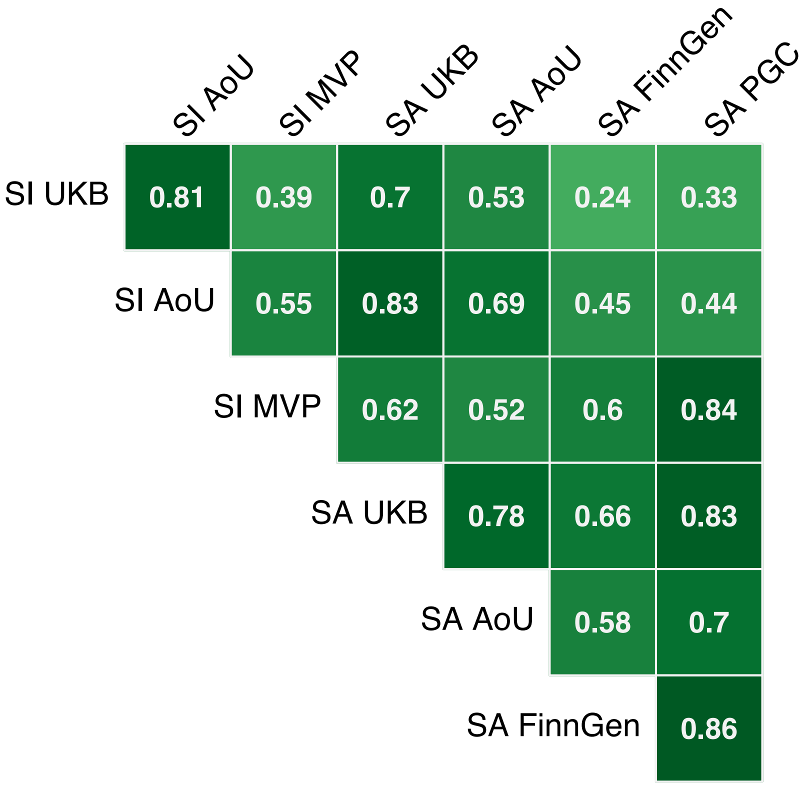


**Figure S1. Genetic correlations of suicide phenotypes across cohorts in European ancestry.** SI: suicide ideation, SA: suicide attempt.

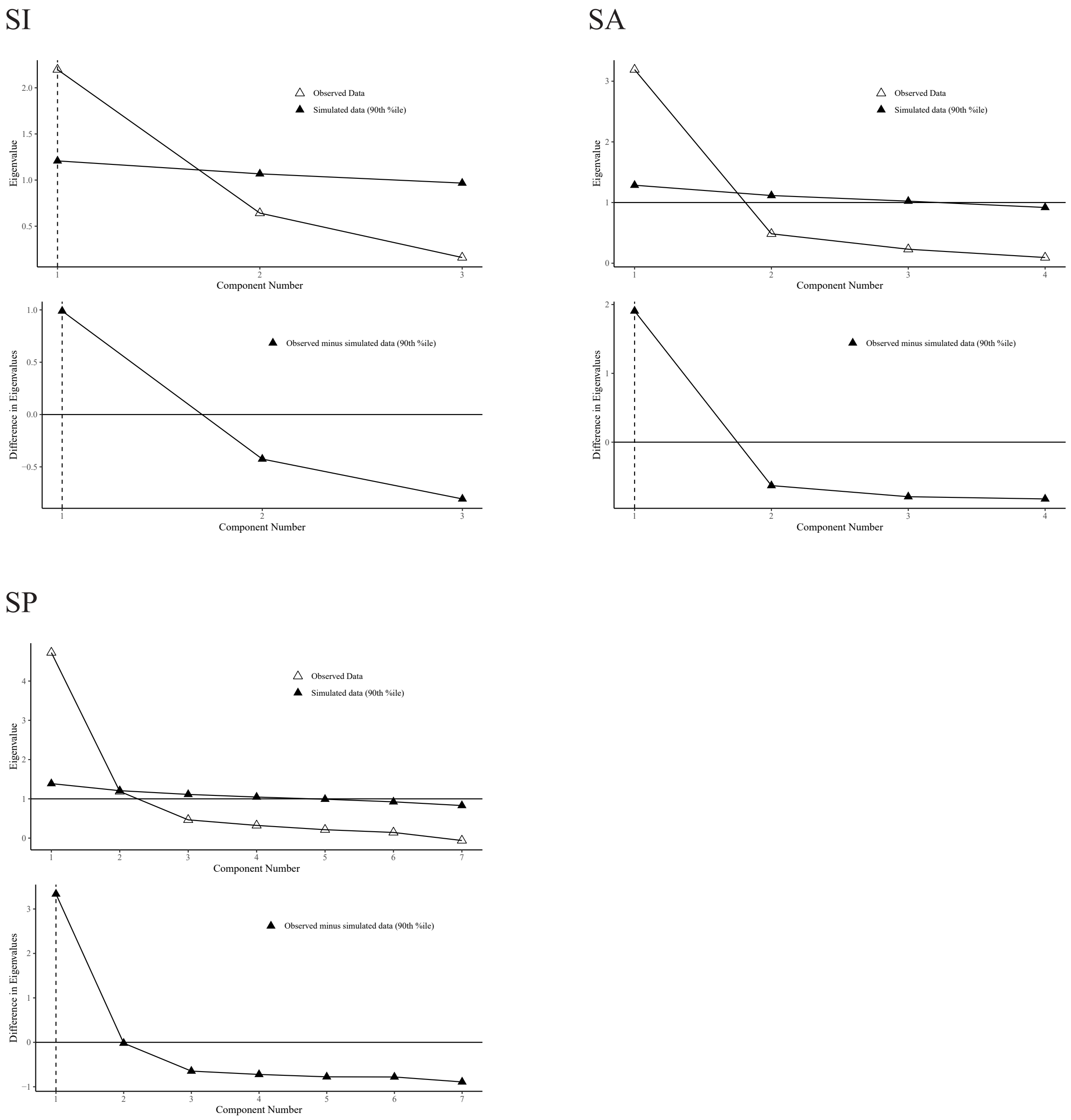


**Figure S2. Parallel analysis (paLDSC) for determining the number of factors for suicide phenotypes in genomic SEM analysis.** SI: suicide ideation, SA: suicide attempt, SP: suicidality spectrum.

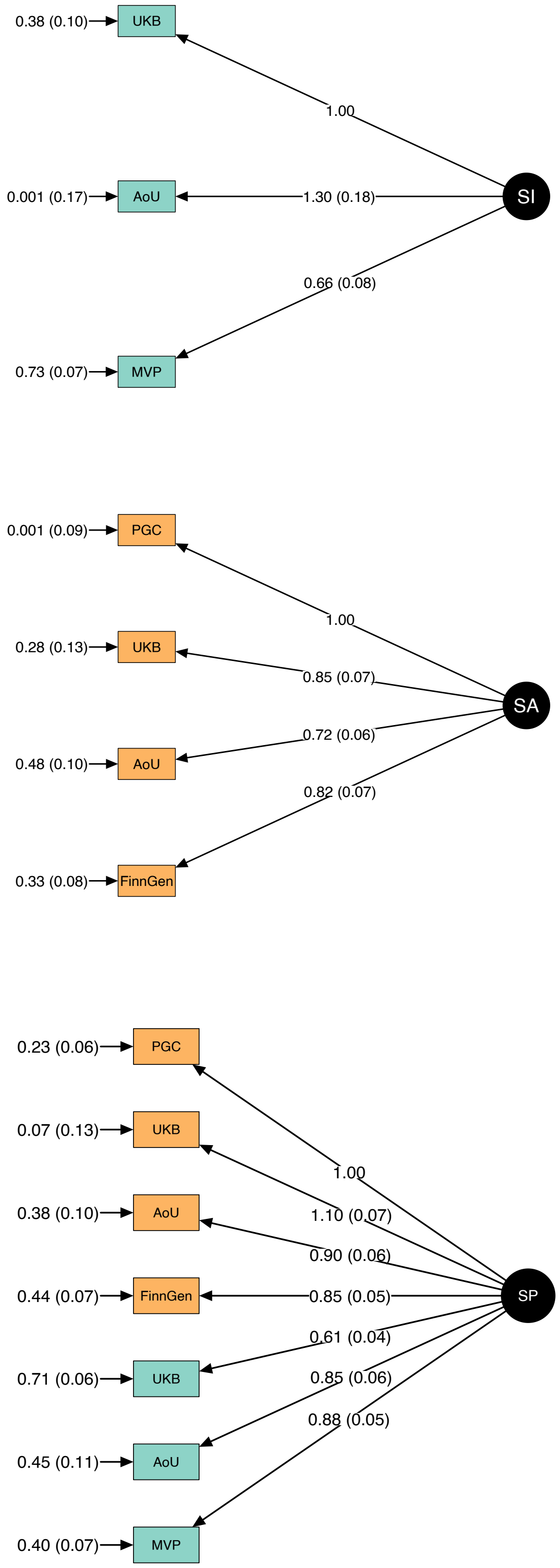


**Figure S3. Factor structure of suicide phenotypes in individuals of European ancestry.** Factor loadings were estimated by confirmatory factor analysis using genomic SEM. SI: suicide ideation, SA: suicide attempt, SP: suicidality spectrum.

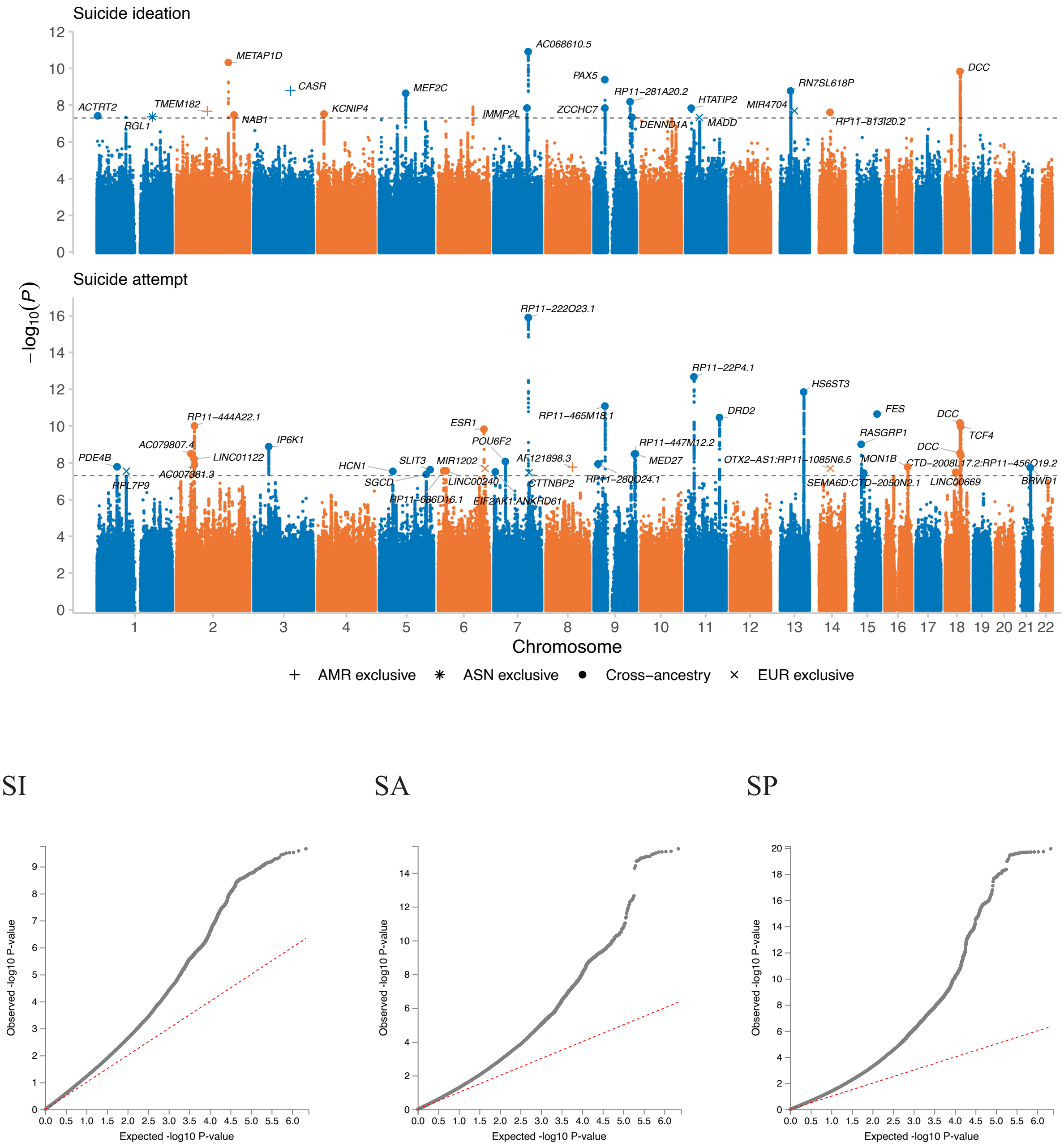


**Figure S4. Manhattan plots and QQ plots of GWAS for suicide phenotypes.** Manhattan plots are based on GWAS effects from the cross-ancestry meta-analysis, with additional independent signals identified within each ancestry. Lead SNPs and their mapped genes are labeled. QQ plots are based on GWAS conducted in European ancestry. SI: suicide ideation, SA: suicide attempt, SP: suicidality spectrum.

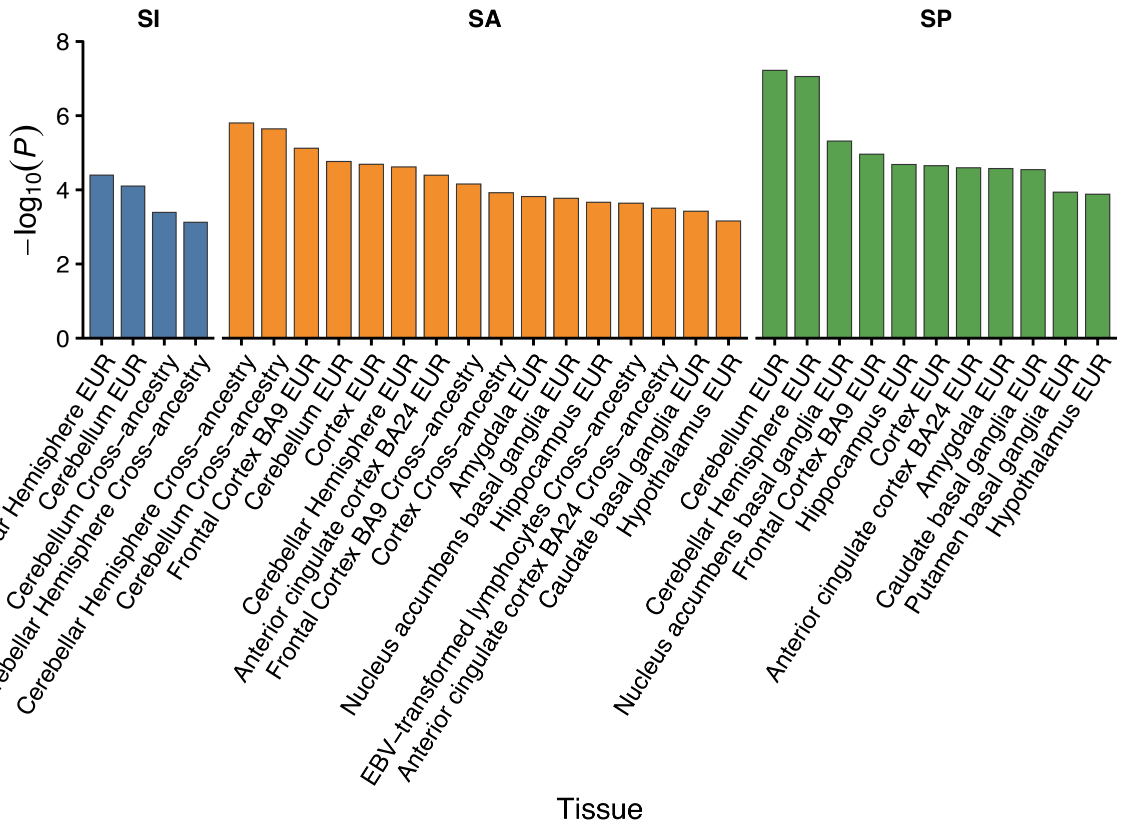


**Figure S5. Significant tissues identified from MAGMA gene expression analysis of suicide phenotypes across ancestries.** SI: suicide ideation, SA: suicide attempt, SP: suicidality spectrum.

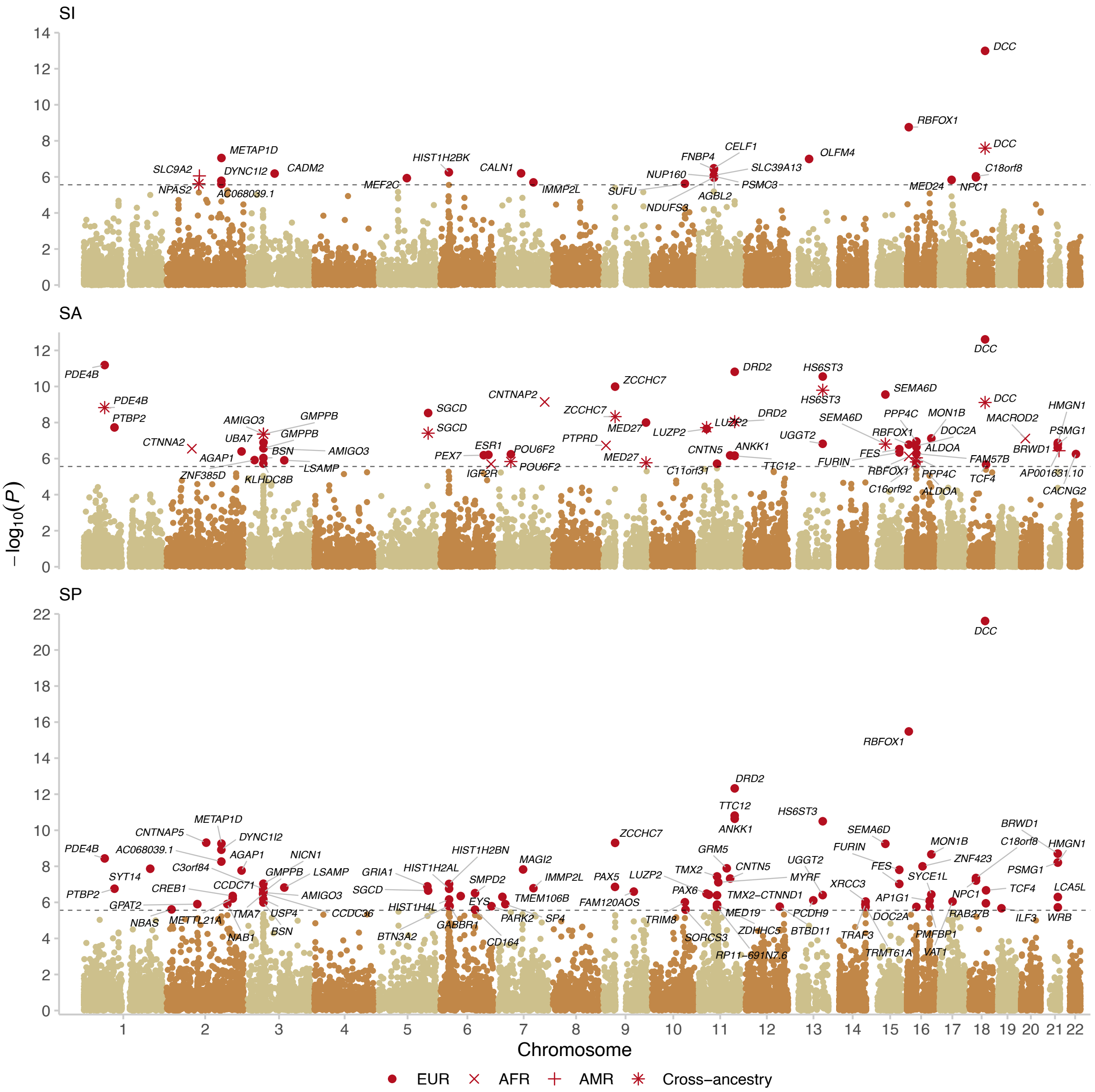


**Figure S6. Manhattan plots of MAGMA gene-based analysis for suicide phenotypes.** Point plots were based on results of European ancestry, with significant genes identified from analyses in other ancestries and cross-ancestry meta-analysis also labeled. SI: suicide ideation, SA: suicide attempt, SP: suicidality spectrum.

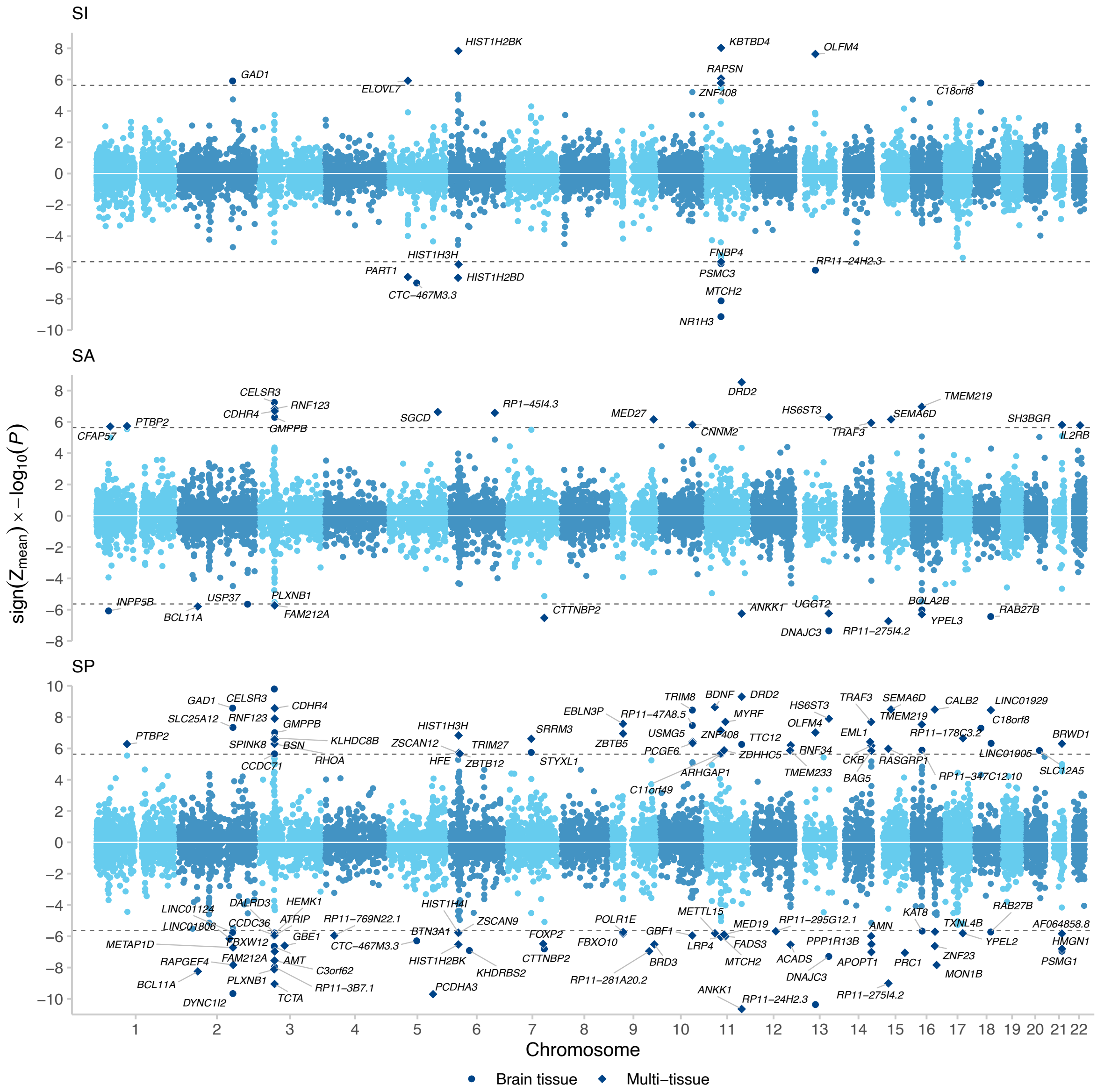


**Figure S7. Manhattan plots of TWAS analysis for suicide phenotypes.** Circles represent genes from multi-tissue analysis using 13 GTEx brain tissues, while diamonds indicate additional significant genes identified when all 49 GTEx tissues were included. SI: suicide ideation, SA: suicide attempt, SP: suicidality spectrum.

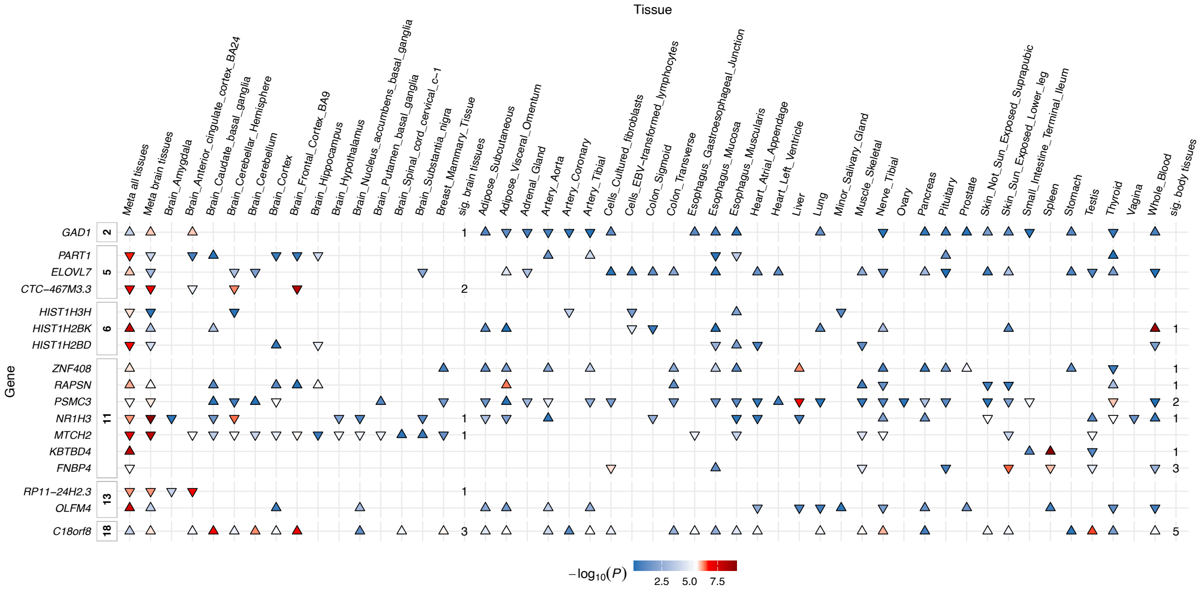


**Figure S8. Effects of genes in multi-tissue and tissue-specific TWAS analyses for suicide ideation.** Only the significant genes identified in multi-tissue analyses using 13 GTEx brain tissues or 49 GTEx tissues are shown in the plot. Pink triangles indicate significant genes in each analyses, with up and down triangles representing positive and negative z-scores, respectively.

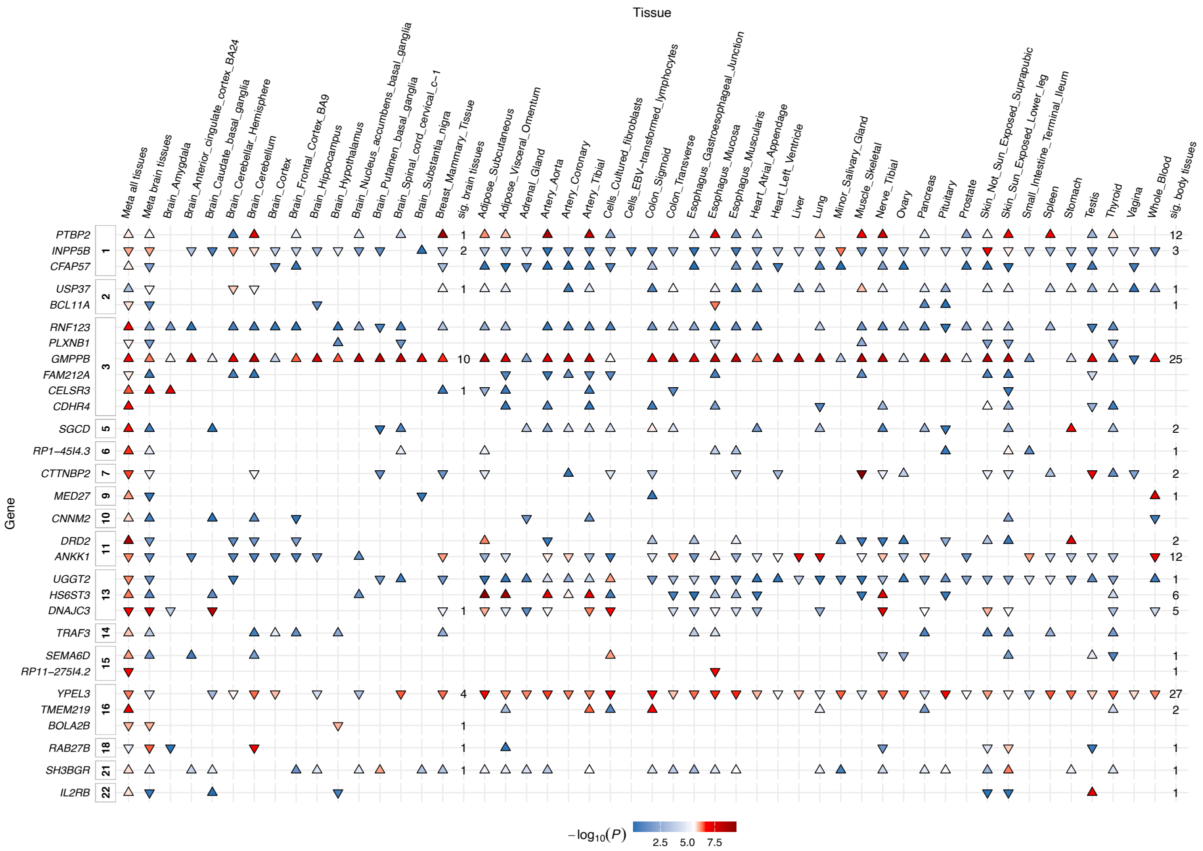


**Figure S9. Effects of genes in multi-tissue and tissue-specific TWAS analyses for suicide attempt.** Only the significant genes identified in multi-tissue analyses using 13 GTEx brain tissues or 49 GTEx tissues are shown in the plot. Pink triangles indicate significant genes in each analyses, with up and down triangles representing positive and negative z-scores, respectively.

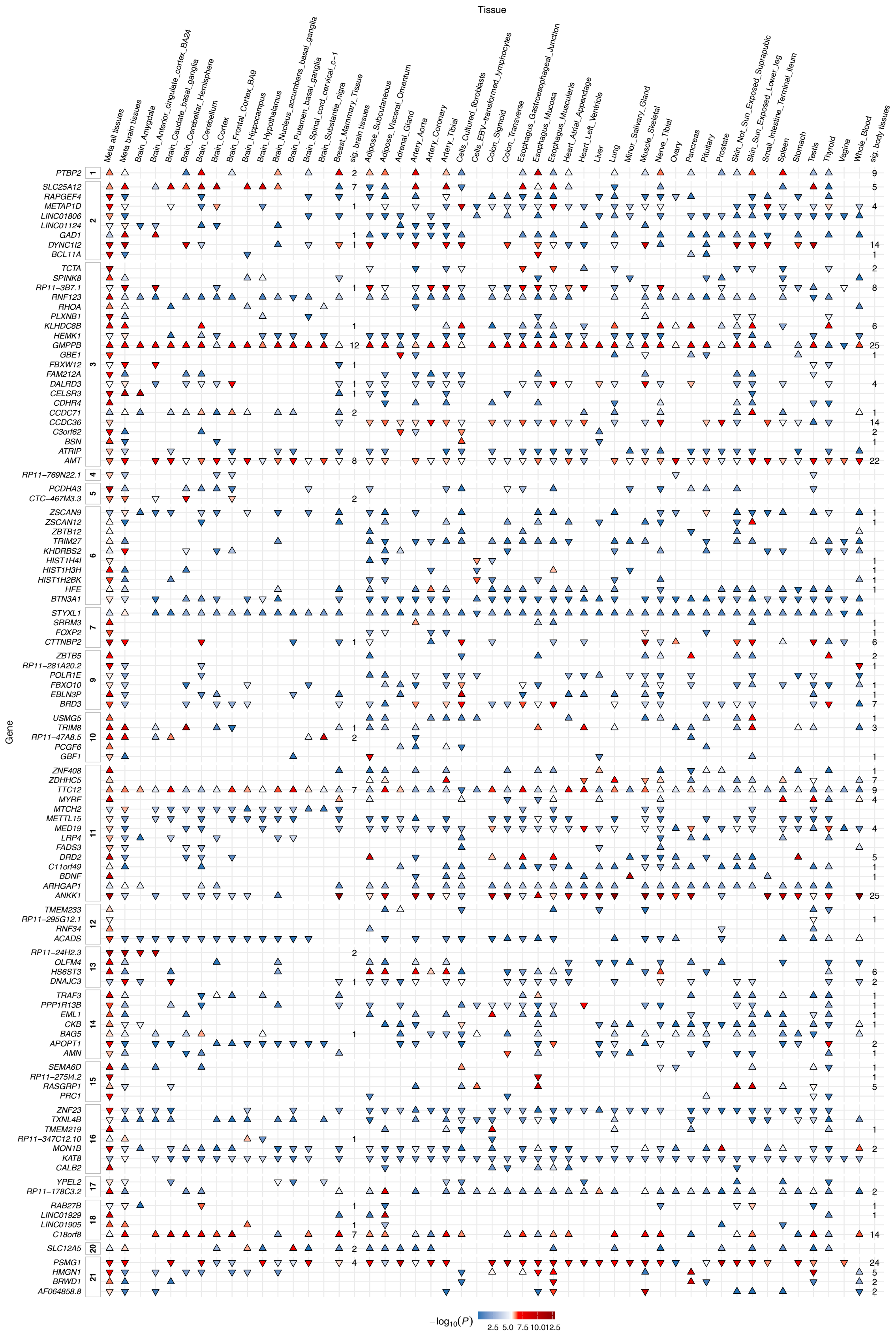

**Figure S10. Effects of genes in multi-tissue and tissue-specific TWAS analyses for suicidality spectrum.** Only the significant genes identified in multi-tissue analyses using 13 GTEx brain tissues or 49 GTEx tissues are shown in the plot. Pink triangles indicate significant genes in each analyses, with up and down triangles representing positive and negative z-scores, respectively.

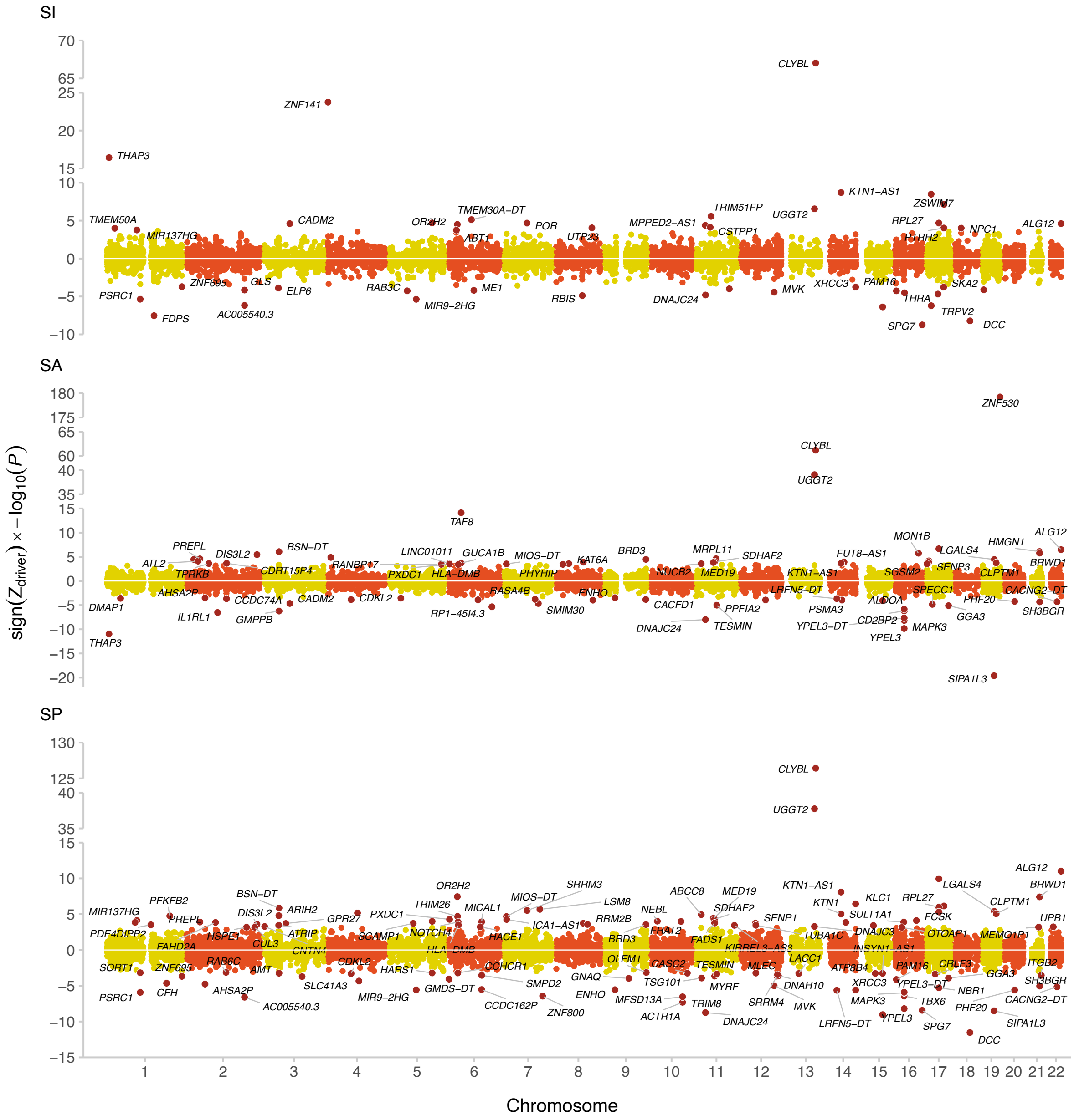


**Figure S11. Manhattan plots of isoform-level TWAS analysis for suicide phenotypes.** Each point on the x-axis represents a gene, accompanied by -log$_{10}$(screening *P*) on the y-axis. Of note, the aggregated Cauchy association test, as utilized by the p_screen function in the isoTWAS package, was used to aggregate isoform-level *P*-values into a gene-level *P*-value, denoted by screening *P*. Significant genes associated with suicide phenotypes are labeled. SI: suicide ideation, SA: suicide attempt, SP: suicidality spectrum.

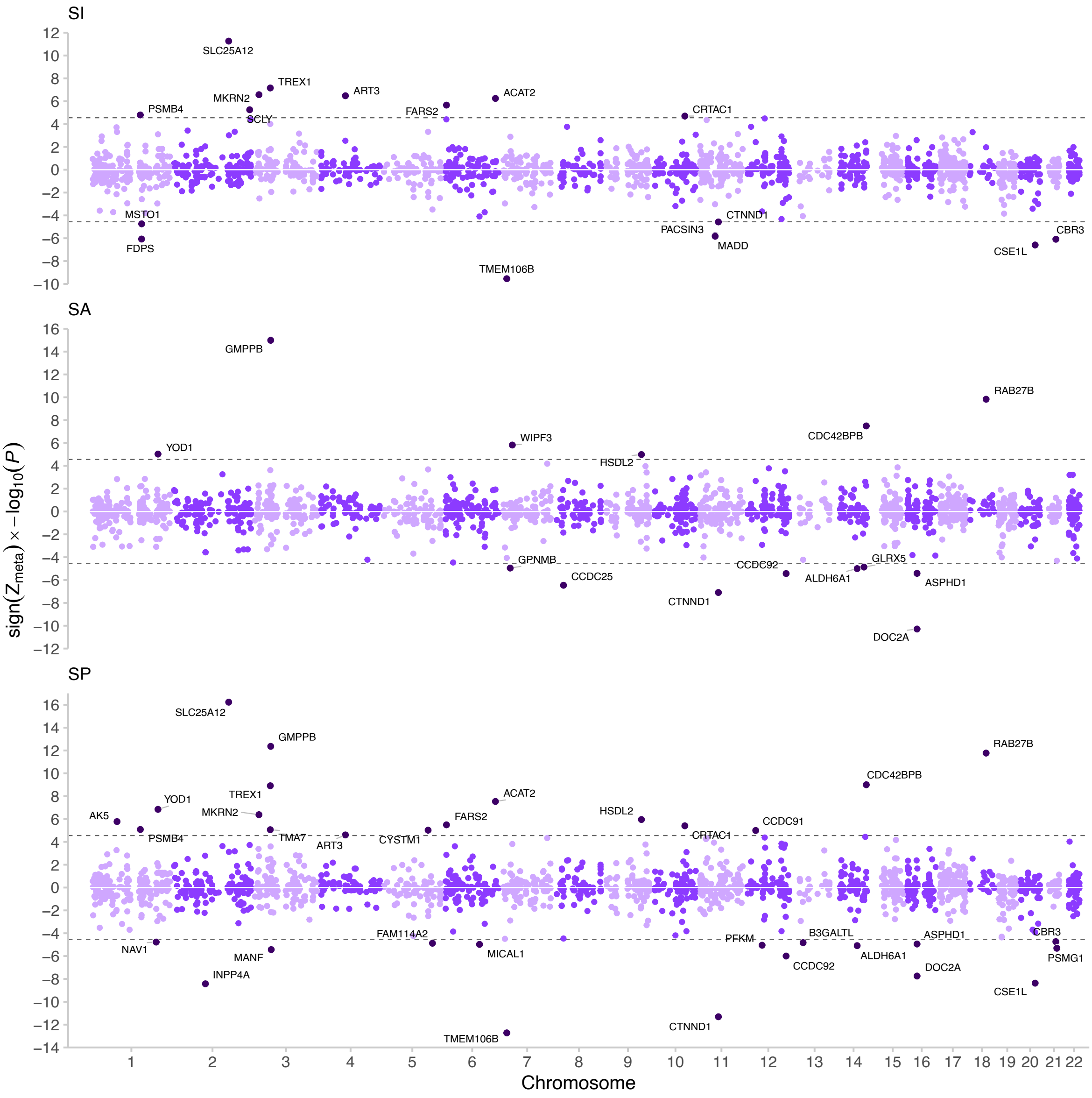


**Figure S12. Manhattan plots of PWAS analysis for suicide phenotypes.** Significant proteins associated with suicide phenotypes are labeled. SI: suicide ideation, SA: suicide attempt, SP: suicidality spectrum.

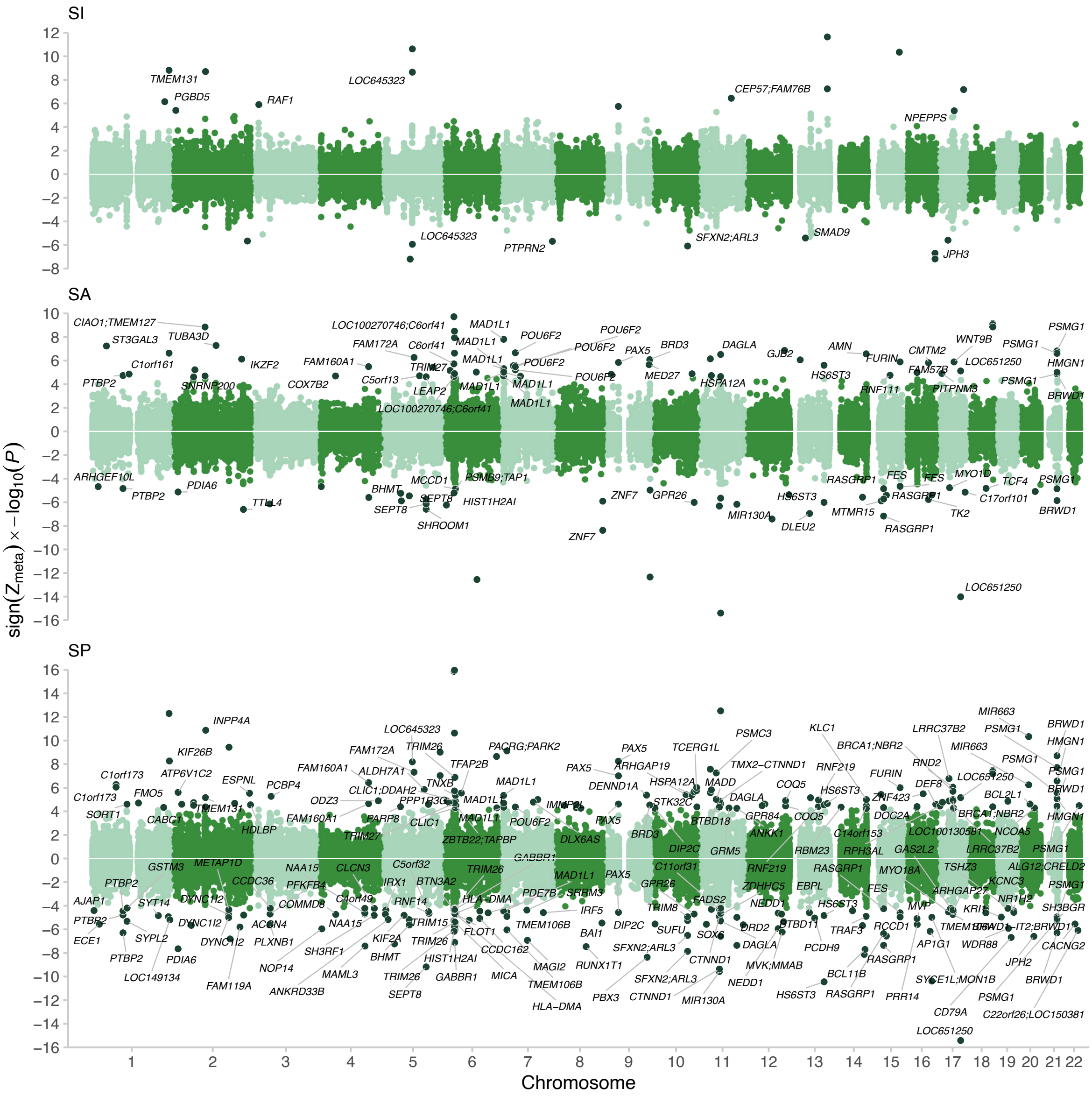


**Figure S13. Manhattan plots of SMR analysis for suicide phenotypes.** Significant CpG sites and their mapped genes are labeled. SI: suicide ideation, SA: suicide attempt, SP: suicidality spectrum.

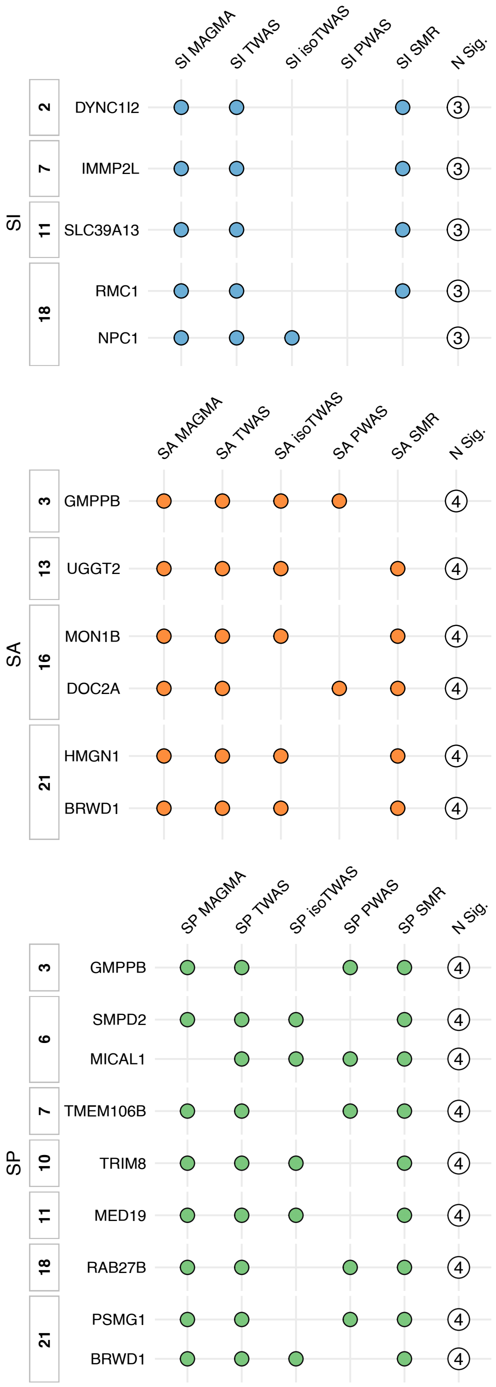


**Figure S14. Top genes associated with each suicide phenotype supported by evidence from five gene discovery analyses.** SI: suicide ideation, SA: suicide attempt, SP: suicidality spectrum.

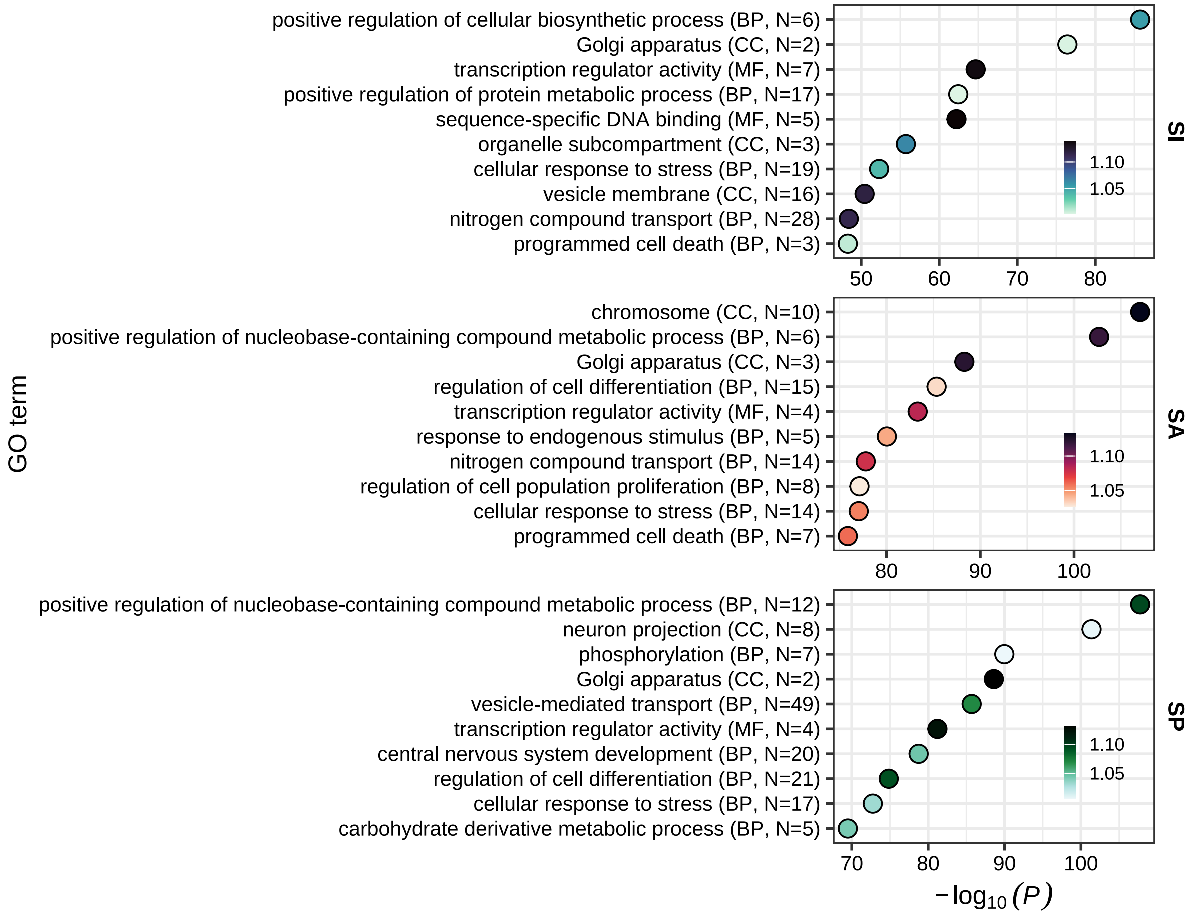


**Figure S15. Reduced Gene Ontology (GO) term groups with significant SNP-based heritability enrichment for suicide phenotypes identified using rrvgo.** BP: Biological Process, CC: Cellular Component, MF: Molecular Function, SI: suicide ideation, SA: suicide attempt, SP: suicidality spectrum. The numbers in parentheses indicate the GO terms included in each group.

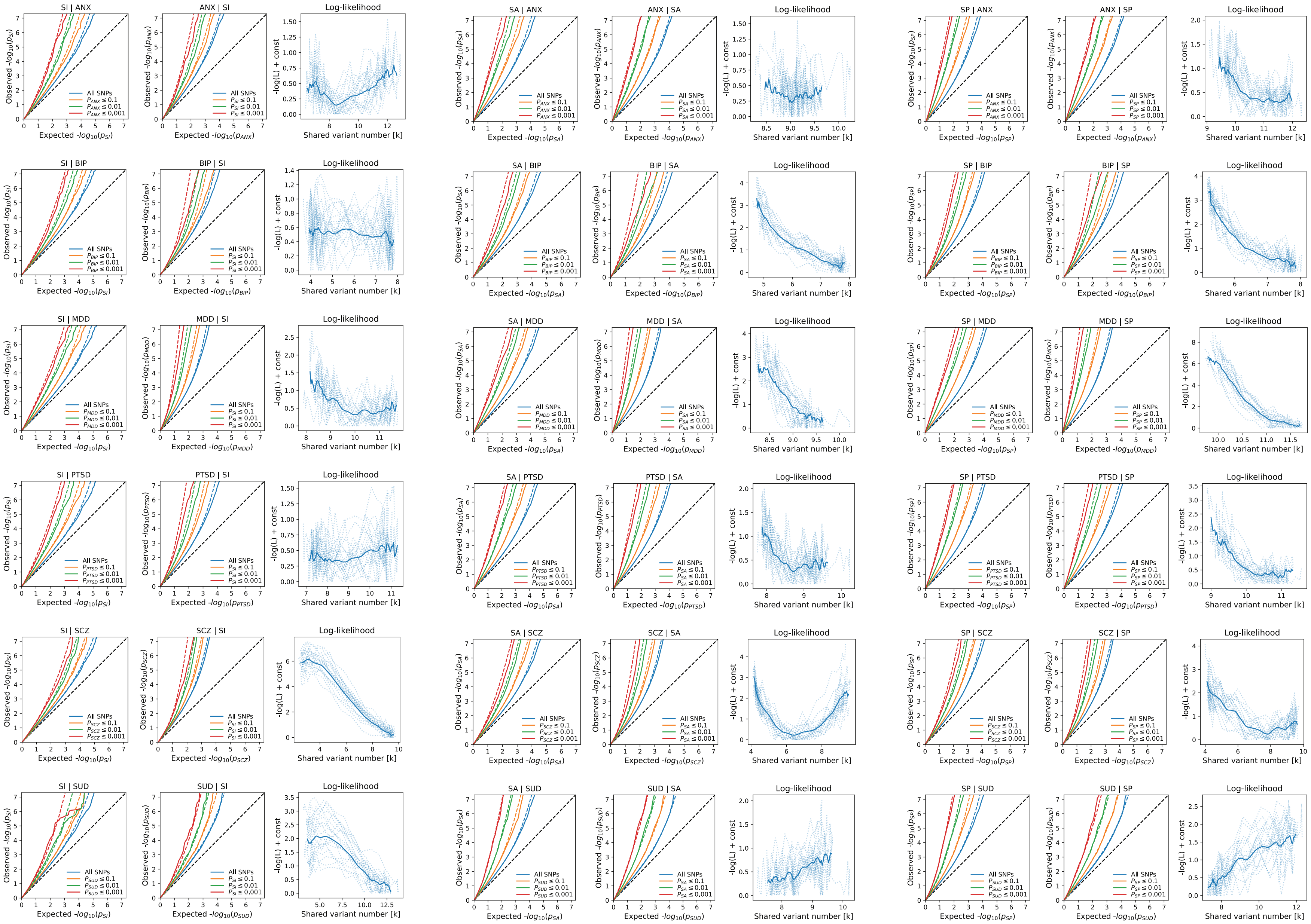


**Figure S16. Conditional QQ plots and log-likelihood plots of MiXeR analysis between suicide phenotypes and psychiatric disorders.** SI: suicide ideation, SA: suicide attempt, SP: suicidality spectrum, MDD: major depressive disorder, PTSD: post-traumatic stress disorder, ANX: anxiety , BIP: bipolar disorder, SCZ: schizophrenia, SUD: substance use disorders.

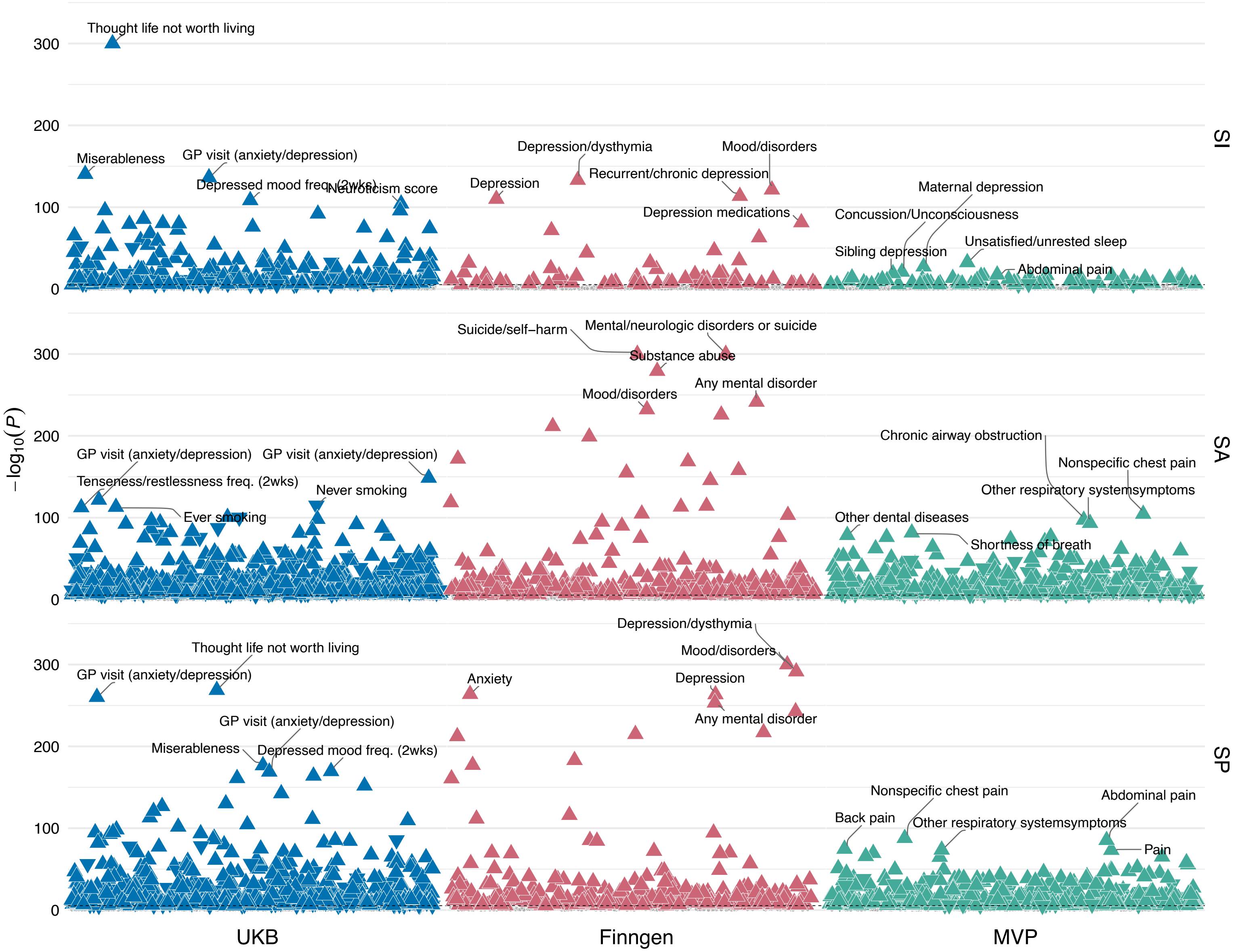


**Figure S17. Phenome-wide genetic correlations between suicide phenotypes and complex traits from UK Biobank (UKB), Finngen, Million Veteran Program (MVP).** SI: suicide ideation, SA: suicide attempt, SP: suicidality spectrum. The top five significant traits in each cohort are labeled.
